# Psychological Distress Before and During the COVID-19 Pandemic Among Adults in the United Kingdom: Coordinated Analyses of 11 Longitudinal Studies

**DOI:** 10.1101/2021.10.22.21265368

**Authors:** Kishan Patel, Elaine Robertson, Alex S. F. Kwong, Gareth J Griffith, Kathryn Willan, Michael J Green, Giorgio Di Gessa, Charlotte F. Huggins, Eoin McElroy, Ellen J. Thompson, Jane Maddock, Claire L Niedzwiedz, Morag Henderson, Marcus Richards, Andrew Steptoe, George B. Ploubidis, Bettina Moltrecht, Charlotte Booth, Emla Fitzsimons, Richard Silverwood, Praveetha Patalay, David Porteous, Srinivasa Vittal Katikireddi

## Abstract

**Importance:** How population mental health has evolved across the COVID-19 pandemic under varied lockdown measures is poorly understood, with impacts on health inequalities unclear.

**Objective:** We investigated changes in mental health and sociodemographic inequalities from before and across the first year of the COVID-19 pandemic in 11 longitudinal studies.

**Design, Setting and Participants:** Data from 11 UK longitudinal population-based studies with pre-pandemic measures of psychological distress were jointly analysed and estimates pooled. Multi-level regression was used to examine changes in psychological distress from pre-pandemic to during the first year of the COVID-19 pandemic.

**Exposures:** Trends in the prevalence of poor mental health were assessed pre-pandemic (TP0) and at three pandemic time periods: initial lockdown (TP1, Mar-June 20); easing of restrictions (TP2, July-Oct 20); and a subsequent lockdown (TP3, Nov 20-Mar 21). We stratified analyses by sex, ethnicity, education, age, and UK country.

**Main Outcomes and Measures:** Psychological distress was assessed using the General Health Questionnaire 12 (GHQ-12), Kessler-6, 9-item Malaise Inventory, Short Mood and Feelings Questionnaire (SMFQ), Patient Health Questionnaire-8 and 9 (PHQ-8/9), Hospital Anxiety and Depression Scale (HADS) and Centre for Epidemiological Studies – Depression (CES-D), across different studies.

**Results:** In total, 49,993 adult participants (61.2% female; 8.7% Non-White) were analysed. Across the 11 studies, mental health deteriorated from pre-pandemic scores across all three pandemic time periods, but with considerable heterogeneity across the study-specific effect sizes estimated (pooled estimate TP1 Standardised Mean Difference (SMD): 0.15 (95% CI: 0.06, 0.25); TP2 SMD: 0.18 (0.09, 0.27); TP3 SMD: 0.21 (0.10, 0.32)). Changes in psychological distress across the pandemic were higher in females (TP3 SMD: 0.23 (0.11, 0.35)) than males (TP3 SMD: 0.16 (0.06, 0.26)), and lower in below-degree level educated persons at TP3 (SMD: 0.18 (0.06, 0.30)) compared to those who held degrees (SMD: 0.26 (0.14, 0.38)). Increased psychological distress was most prominent amongst adults aged 25-34 and 35-44 years compared to other age groups. We did not find evidence of changes in distress differing by ethnicity or UK country.

**Conclusions and Relevance:** The substantial deterioration in mental health seen in the UK during the first lockdown did not reverse when lockdown lifted, and a sustained worsening was observed across the pandemic. Mental health declines have been unequal across the population, with females, those with higher degrees, and those aged 25-44 years more affected.

## Introduction

There have been widespread concerns about the impact of the COVID-19 pandemic and related mitigation measures on population mental health.^1,2^ Globally, there is evidence that the pandemic has resulted in poorer mental health,^3^ but much of this might depend on COVID-19 rates and the varying mitigation policies implemented. Concerns exist that specific policy responses, notably so-called ‘lockdown’ measures, may themselves adversely impact mental health. Examining changes from before the pandemic, but also across different pandemic periods with different restrictions in place, may help understand the drivers of mental health impacts.

Reports on population mental health changes at the start of the pandemic within the UK are conflicting, with some studies indicating a wide-spread decline in psychological wellbeing early on,^4^ while other studies suggest improvements or no changes in mental ill-health.^5,6^ Findings have remained inconsistent as the pandemic has progressed, with both increasing and decreasing levels of poorer mental health reported.^7–9^

The COVID-19 pandemic has had disproportionate impacts on different age and socio-demographic groups via different mechanisms.^10,11^ For instance, older adults were at greater risk of severe disease and were asked to stay at home and minimise face-to-face contact (shielding), whilst younger people, women, and ethnic minorities have been disproportionately impacted by employment losses and precarity.^12^ The focus of many existing studies is on population averages which may have concealed inequalities in impacts on mental health.^3^

Uncertainty remains about how mental health has changed over the pandemic, including who has been most affected and whether any observed deterioration reflects lockdown measures or other aspects of the pandemic. To examine this, we conducted co-ordinated analyses of 11 UK longitudinal population studies with data from before and across the pandemic. We aimed to: 1) estimate the impacts of the pandemic on population mental health and how these evolved during the first year of the pandemic as lockdown restrictions changed; and 2) examine inequalities in these impacts by age, sex, ethnicity, education level, and UK country.

## Methods

### Design

The UK National Core Studies – Longitudinal Health and Wellbeing initiative aims to co-ordinate primary analyses across multiple UK longitudinal population-based studies.^13,14^ Co-ordinating analyses across different datasets minimises methodological heterogeneity and maximises comparability, while appropriately accounting for the study design and characteristics of individual datasets.

### Participants

Data were pooled from 11 UK longitudinal population studies which conducted surveys both before and during the COVID-19 pandemic. Details of the design, sampling frames, current age range, timing of the pre-pandemic and COVID-19 surveys, response rates, and analytical sample size are in Table 1, with further details of each analytical sample in Supplement 1.

**Table 1.** Details of each included study

| Study Population | Design and Sample Frame | 2020 Age Range in years | Most Recent Pre-Pandemic Survey | Details of COVID-19 Surveys (Response Rate) | Mental Distress Measure Used | Analytic N |
| --- | --- | --- | --- | --- | --- | --- |
| *Age Homogenous Cohorts* |  |  |  |  |  |  |
| MCS: Millennium Cohort Study<sup>15</sup> | Cohort of UK children born between Sept 2000 and Jan 2002 with regular follow-up surveys from birth. | 18-20 | 2018 | Three surveys: May (26.6%); Sep-Oct (24.2%); Feb-Mar (22%) | Kessler-6 (K6)<sup>44</sup> | 4,988 |
| ALSPAC: Avon Longitudinal Study of Parents and Children-Generation 1<sup>16</sup> | Cohort of children born in the South-West of England between April 1991 and Dec 1992, with regular follow-up questionnaires from birth. | 27-29 | 2017-2018 | Three surveys: April (19%); June (17.4%); December (26.4%) | Short Mood and Feelings Questionnaire (SMFQ)<sup>45</sup> | 3,208 |
| NS: Next Steps, formerly known as Longitudinal Study of Young People in England<sup>17</sup> | Sample recruited via secondary schools in England at around age 13 with regular follow-up surveys thereafter. | 29-31 | 2015 | Three surveys: May (20.3%); Sep-Oct (31.8%); Feb-Mar (29%) | General Health Questionnaire 12 (GHQ-12)<sup>46</sup> | 4,139 |
| BCS70: British Cohort Study 1970<sup>18</sup> | Cohort of all children born in Great Britain (i.e., England, Wales & Scotland) in one week in 1970, with regular follow-up surveys from birth. | 50 | 2016 | Three surveys: May (40.4%); Sep-Oct (43.9%); Feb-Mar (40%) | 9-item Malaise inventory<sup>47</sup> | 5,532 |
| NCDS: National Child Development Study<sup>19</sup> | Cohort of all children born in Great Britain (i.e., England, Wales & Scotland) in one week in 1958, with regular follow-up surveys from birth. | 62 | 2013 | Three surveys: May (57.9%); Sep-Oct (53.9%); Feb-Mar (52%) | 9-item Malaise inventory<sup>47</sup> | 6,667 |
| NSHD: National Survey of Health and Development<sup>20</sup> | Cohort of all children born in Great Britain (i.e., England, Wales & Scotland) in one week in 1946, with regular follow-up surveys from birth. | 74 | 2015 | Three surveys: May (68.2%); Sep-Oct (61.5%); Feb-Mar (90%) | General Health Questionnaire 12 (GHQ-12)<sup>46</sup> | 2,007 |
| *Age Heterogeneous Studies* |  |  |  |  |  |  |
| USoc: Understanding Society: the UK Household Longitudinal Survey<sup>21</sup> | A nationally representative longitudinal household panel study, based on a clustered-stratified probability sample of UK households, with all adults aged 16+ in chosen households surveyed annually. | 16-96 | 2018-2019 | Seven surveys: April (40.3%); May (33.6%); Jun (32.0%); July (31.2%); Sep (29.2%); Nov (27.3%); | General Health Questionnaire 12 (GHQ-12)<sup>46</sup> | 12,437 |
|  |  |  |  | Jan 2021 (27.2%) |  |  |
| ELSA: English Longitudinal Study of Aging<sup>25</sup> | A nationally representative population study of individuals aged 50+ living in England, with biennial surveys and periodic refreshing of the sample to maintain representativeness. | 52-90+ | 2018-2019 | Two surveys:<br>Jun-July (75%); Nov-Dec (73%) | Centre for Epidemiological Studies – Depression (CES-D)<sup>48</sup> | 5,699 |
| GS: Generation Scotland: The Scottish Family Health Study<sup>22</sup> | A family-structured, population-based Scottish cohort, with participants aged 18-99 recruited between 2006-2011 | 27-100 | 2006-2011 | Three surveys:<br>April-Jun 2020 (21.3%);<br>Jul-Aug 2020 (15.4%);<br>Feb 2021 (14.3%) | Patient Health Questionnaire 9 (PHQ-9)<sup>49</sup> or Patient Health Questionnaire 8 (PHQ-8)<sup>50</sup> and Generalised Anxiety Disorder Assessment 7 (GAD-7)<sup>51</sup> | 4,151 |
| TwinsUK: the UK Adult Twin Registry<sup>23</sup> | A cohort of volunteer adult TwinsUK (55% monozygotic and 43% dizygotic) from around the United Kingdom who were sampled between 18-101 years of age. | 22-96 | 2017-2018 | Three surveys:<br>April (64.3%); July (77.6%); November (76.1%) | Hospital and Anxiety Depression Scale (HADS)<sup>52</sup> | 4,040 |
| BiB: Born in Bradford<sup>24</sup> | Two birth cohorts recruiting pregnant women and their children between 2007 and 2010 (BiB Growing Up), and from 2016 (Born in Bradford's Better Start (BiBBS)) | 16-57 | 2016-2020 | Two surveys:<br>April-Jun (28%); Oct-Nov (35.8%) | Patient Health Questionnaire 9 (PHQ-9)<sup>49</sup> or Patient Health Questionnaire 8 (PHQ-8)<sup>50</sup> and Generalised Anxiety Disorder Assessment 7 (GAD-7)<sup>51</sup> | 1,967 |

Six studies were age homogenous cohorts (similar aged individuals): Millennium Cohort Study (MCS);^15^ Children in the Avon Longitudinal Study of Parents and Children (ALSPAC);^16^ Next Steps (NS; formerly known as the Longitudinal Study of Young People in England);^17^ 1970 British Cohort Study (BCS70);^18^ 1958 National Child Development Study (NCDS);^19^ and 1946 National Survey of Health and Development (NSHD).^20^ Five other studies had age heterogeneous samples (i.e. cohorts with multiple age groups): Understanding Society (USoc);^21^ Generation Scotland (GS);^22^ Twins UK (TwinsUK);^23^ Born in Bradford (BiB);^24^ and the English Longitudinal Study of Ageing (ELSA).^25^

Analytical samples included those who had valid observations of psychological distress in a pre-pandemic survey, at least one survey during the pandemic, and valid data on sex and age (participant flow diagrams for each study in Supplement 1). Participants who had died or emigrated by the start of the pandemic are also excluded. Most studies were weighted to be representative of their target population, accounting for sampling design and differential non-response to the COVID-19 surveys.^26–28^ Weights were not used for ALSPAC, TwinsUK, GS and BiB.

Ethical approvals were received for all included studies, with ethics statements described in Supplement 1.

#### Measures

Below we describe the variables used for analysis. Details of the specific scales and coding used within each cohort are in Supplement 1.

#### Mental Health

Psychological distress was measured both before the pandemic (a range of 0-7 years prior) and at multiple time-points across the pandemic using validated continuous scales measuring symptoms of common mental health disorders such as depression and anxiety (specific measures used in Table 1). Continuous scales were standardised across time-points and within studies on a common standard deviation-based scale. This enhances comparability of estimates between studies while allowing examination of changes over time within studies. We also conducted analyses with dichotomous indicators of high psychological distress using established thresholds for each scale (Supplement 1 for thresholds).

While most studies utilised the same measure for both pre-pandemic and COVID-19 surveys, GS and NSHD used different measures. For these studies, we identified comparable items to create a smaller scale consistent over time and the threshold for the binary outcome was re-weighted based on the number of items retained (Supplement 1 for details).

#### Pandemic Time Period

We identified three time periods (TP1-TP3) representing different stages over the course of the pandemic in the UK, for comparison against pre-pandemic mental health (TP0). Surveys from April-June 2020 represented the first wave of high infection levels accompanied by the first ‘lockdown’ measures (TP1). Surveys taken from July-October 2020 coincided with easing of restrictions and lower rates of infection (TP2). Following this, infection levels again increased, and lockdown measures were reintroduced; surveys taken from November 2020-March 2021 represent this second wave of infections (TP3). Some studies contributed multiple survey waves to some time periods and not all studies were represented in all three COVID-19 time periods (Table 1).

#### Covariates

The following covariates were adjusted for and/or used to stratify estimates: sex (male; female); age in the age heterogeneous cohorts (coded in 10-year bands to examine non-linearity: 16-24, 25-34, 35-44, 45-54, 55-64, 65-74, 75+); ethnicity (self-reported and coded for main analyses; as White-including White ethnic minorities vs. Non-White ethnic minorities); UK country of residence (England, Scotland, Wales or Northern Ireland); and highest educational qualification (degree vs. less than degree; parental education was used for the MCS cohort, who had not all completed their full-time education).

### Statistical Analysis

Changes in continuous measure of mental health over the three time periods were modelled using multi-level mixed effects models within each study to account for correlations between repeated measures from the same individuals, adjusting for sex and age (in age heterogeneous cohorts). Time period was a categorical exposure, with TP0 as the reference. In some studies, multiple survey waves were included within the same time period. Coefficients are presented as standardised mean differences (SMD). Multi-level mixed effects Poisson regression models with robust standard errors were used to calculate relative risks for the binary outcome.^29^

Results from each study were pooled using a random effects meta-analysis with restricted maximum likelihood. Meta-analyses were conducted separately for continuous psychological distress scores, and binary high psychological distress thresholds. Heterogeneity is reported using the I^2^ statistic.^30^ Interactions between time period and sex, education and ethnicity were estimated within each study and then meta-analysed to formally test for effect modification (i.e., to determine whether changes across time periods varied between population subgroups). Formal interactions could not be tested by age and UK country, given the age homogeneous nature of several cohorts and few studies included all UK nations. We present meta-analysis of estimates stratified by sex, education, ethnicity, age and UK country.

Further sensitivity meta-analyses restricted analyses to include studies that only assessed anxiety specifically, assessed depression specifically, and that included survey responses for all three time periods. To explore the heterogeneity in estimates, meta-regression analyses were conducted, quantifying the effects of time between pre- and post-pandemic measures, measurement type, and whether study samples were representative of their target age range in the UK population (Supplement 1 for studies included in each analysis). All meta-analyses and meta-regressions were conducted using Stata 17 (StataCorp LP).

## Results

Across 11 individual longitudinal studies, 49,993 participants were analysed, ranging from 1,816 participants in NSHD to 12,437 in USOC. The proportion of females ranged between 50.0% in NCDS to 100.0% for BiB and ethnic minorities ranged from 0.6% in GS and TwinsUK to 62.2% in BiB. Descriptive statistics for all the studies, weighted and taking account of complex survey design where relevant, are in Supplement 1.

### Descriptive analysis

Figure 1a shows that for most studies, prevalence of high psychological distress either worsened or was fairly stable over the course of the pandemic. The largest increase in prevalence of high psychological distress was observed within the ELSA study, rising from 11.5% to 28.0% over the course of the three time periods. The largest increase between two consecutive time periods was observed within the NSHD study, between the pre-pandemic (2015) and first pandemic time periods, rising from 11.4% to 35.0%. In two studies (ALSPAC and BCS70), the prevalence of distress in the final pandemic time period (TP3) was marginally lower than the pre-pandemic time period (prevalence reduced by 2.3% and 0.8% respectively).

**Figure 1.**
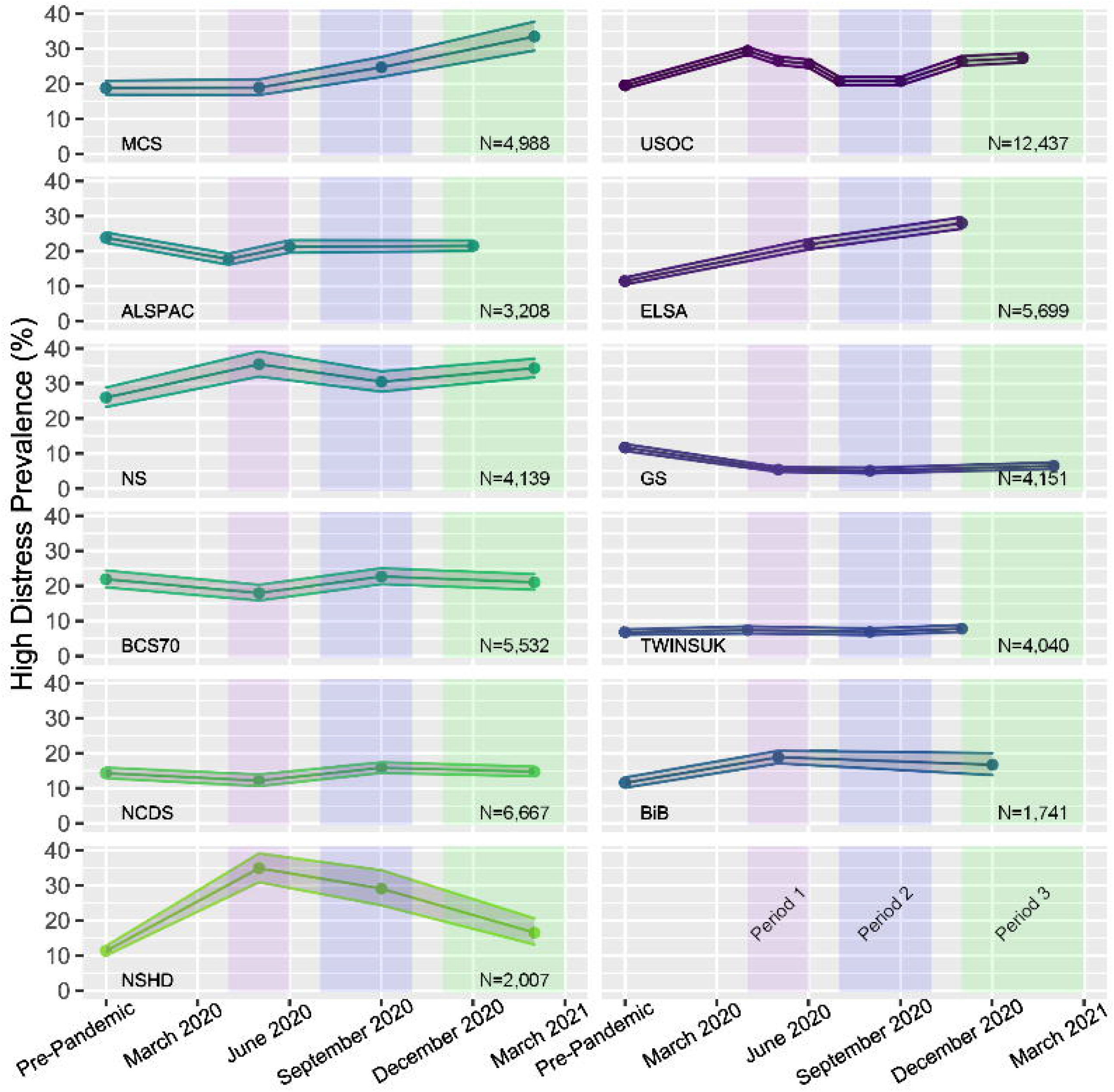
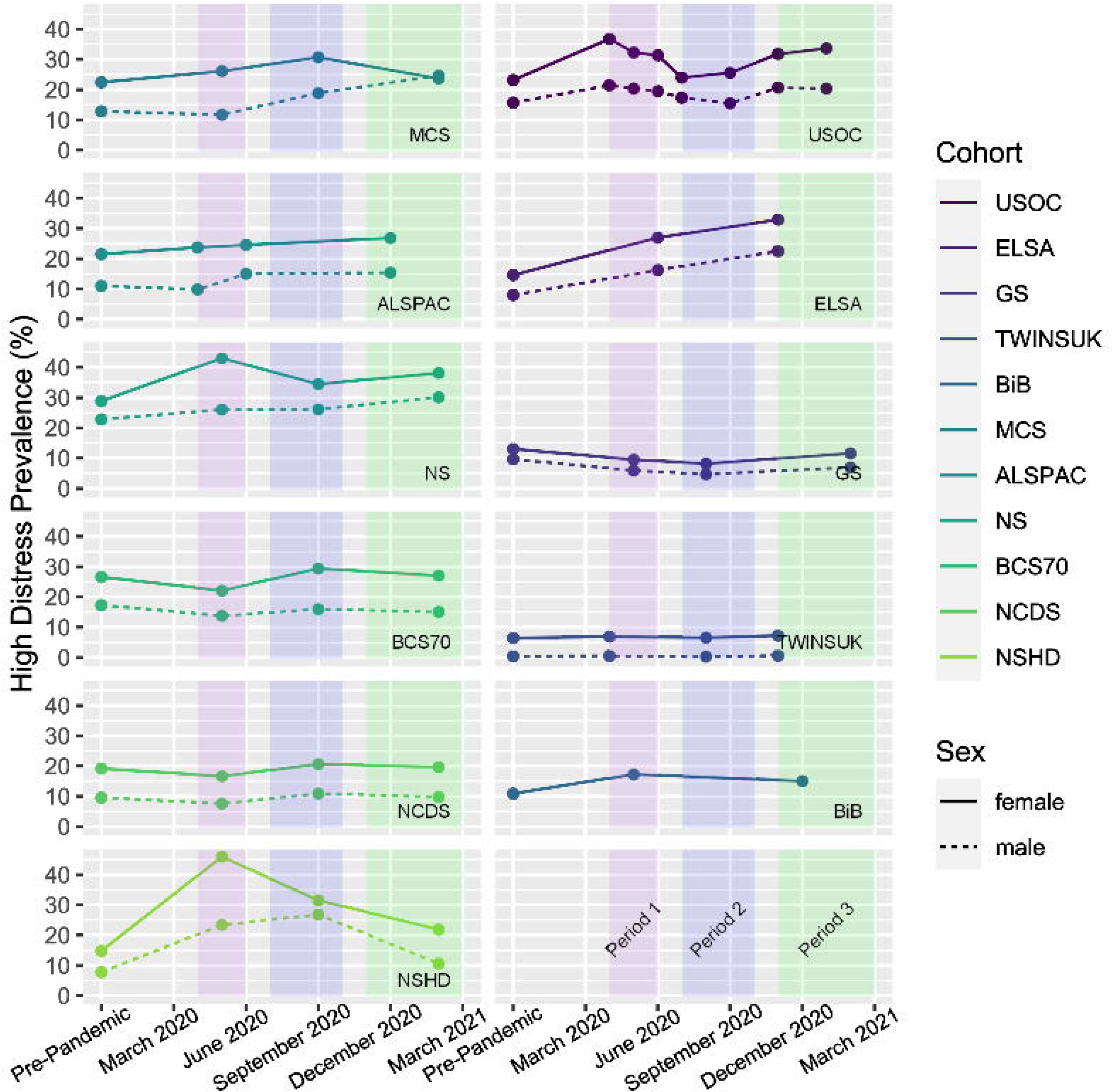
Trends in overall prevalence (1a) and sex stratified prevalence (1b) estimates of high psychological distress, within each of the 11 longitudinal studies analysed in this paper, as defined by measure-specific thresholds. Coloured boxes indicate the time period groupings used for analysis in this paper.

Figure 1b shows the sex difference in mental health over the course of the pandemic, with higher prevalence of distress among women than men in all sex-heterogeneous studies. In April and May 2020 (TP1), sex inequalities appeared especially high, with female respondents exhibiting higher prevalence of mental distress in most studies.

### Changes in distress from before and during the pandemic: pooled analysis

Psychological distress increased from pre-pandemic scores across all three pandemic time periods examined (observed in eight out of the 11 included cohorts when focussing on general distress or depressive symptom measures), with no clear differences in impacts across the three pandemic time periods (TP1 SMD: 0.15 (95% CI: 0.06, 0.25); TP2 SMD: 0.18 (0.09, 0.27); TP3 SMD: 0.21 (0.10, 0.32)).

However, there was considerable heterogeneity between estimates from different studies (I^2^=99.2%, 98.6%, and 99.2% respectively at 3 TPs), with estimates for TP1 ranging from SMD: −0.08 (−0.11, −0.05) for ALSPAC, to SMD: 0.46 (0.37, 0.55) for NSHD (individual cohort results in Supplement 2). Leave one out meta-analysis revealed that no single cohort significantly skewed the pooled estimates (Supplement 2). Similar patterns and high levels of heterogeneity are observed when considering prevalence of psychological distress as a binary outcome (Supplementary Files 1 and 2). Estimates for both continuous and binary measures of mental distress are displayed in Figure 2.

**Figure 2.**
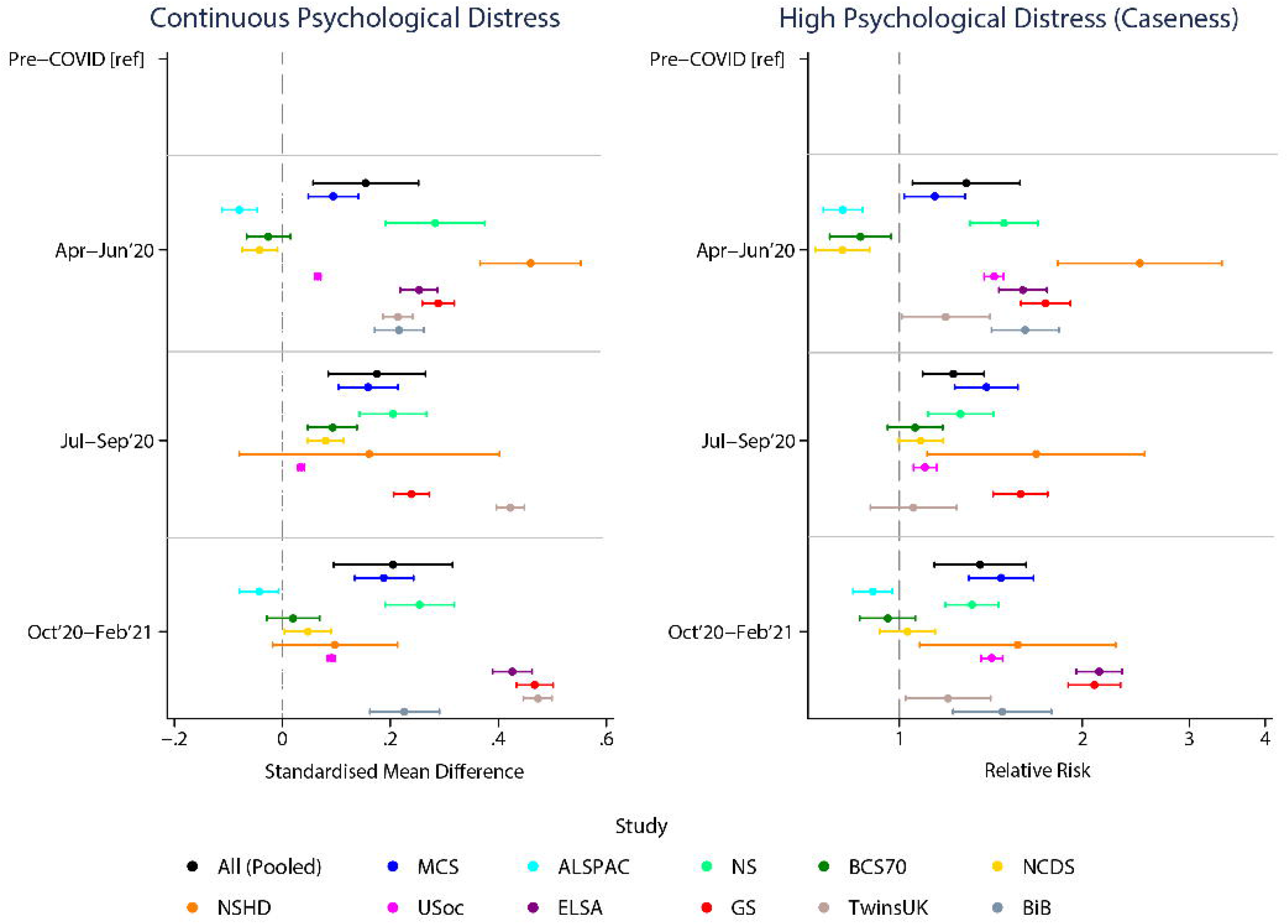
Changes in psychological distress before and during the pandemic in each of 11 longitudinal UK studies (and pooled estimates). Overall, impacts across time periods (compared to pre pandemic distress) for the continuous psychological distress scores (left panel) and binary high distress scores (right panel).

### Inequalities in changes over time periods: pooled analysis

Meta-analysis of the study-specific interaction terms between each marker of inequity and time period indicated that changes in distress were greater in females (TP3 SMD: 0.23 (0.11, 0.35)) compared to males (TP3 SMD: 0.16 (0.06, 0.26)) (Supplement 2) suggesting a further widening of sex inequalities. Changes were marginally lower at TP1 and TP3 for persons with a below-degree level education (TP3 SMD: 0.18 (0.06, 0.30)) compared to those with a degree (TP3 SMD: 0.26 (0.14, 0.38)), albeit often from a greater pre-pandemic inequality, indicating a slight narrowing of educational inequalities during the pandemic. We did not find evidence for trends differing by ethnicity and UK country. Heterogeneity varied across these analyses, with I^2^ values ranging from 44.2% for the interaction between education and TP1, and 88.8% for ethnicity and TP1. Estimates stratified by sex, ethnicity, education, and UK country are shown in Figure 3. Again, in all analyses there was large heterogeneity between study estimates (Supplement 2).

**Figure 3.**
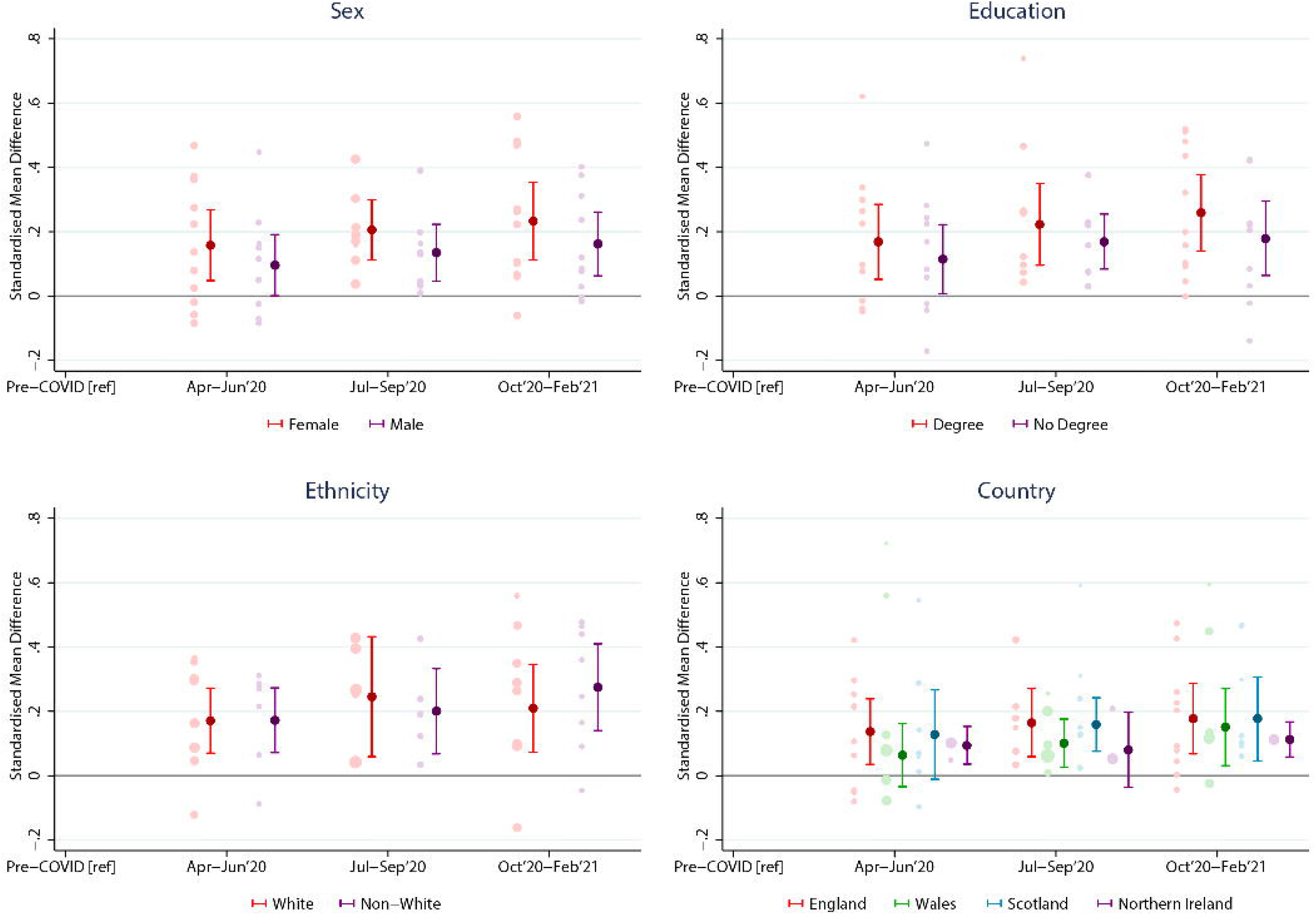
Changes in psychological distress over time by: a) sex; b) education; c) ethnicity; d) UK country. Stratified changes across time periods (compared to pre-pandemic distress). Each light-coloured point represents estimates from a different included study (study specific estimates can be seen in Supplement 2).

Age-stratified results show no monotonic pattern by age (Figure 4), despite some suggestion that the impact of the pandemic on mental health might have been greater in those aged 25-44 years.

**Figure 4.**
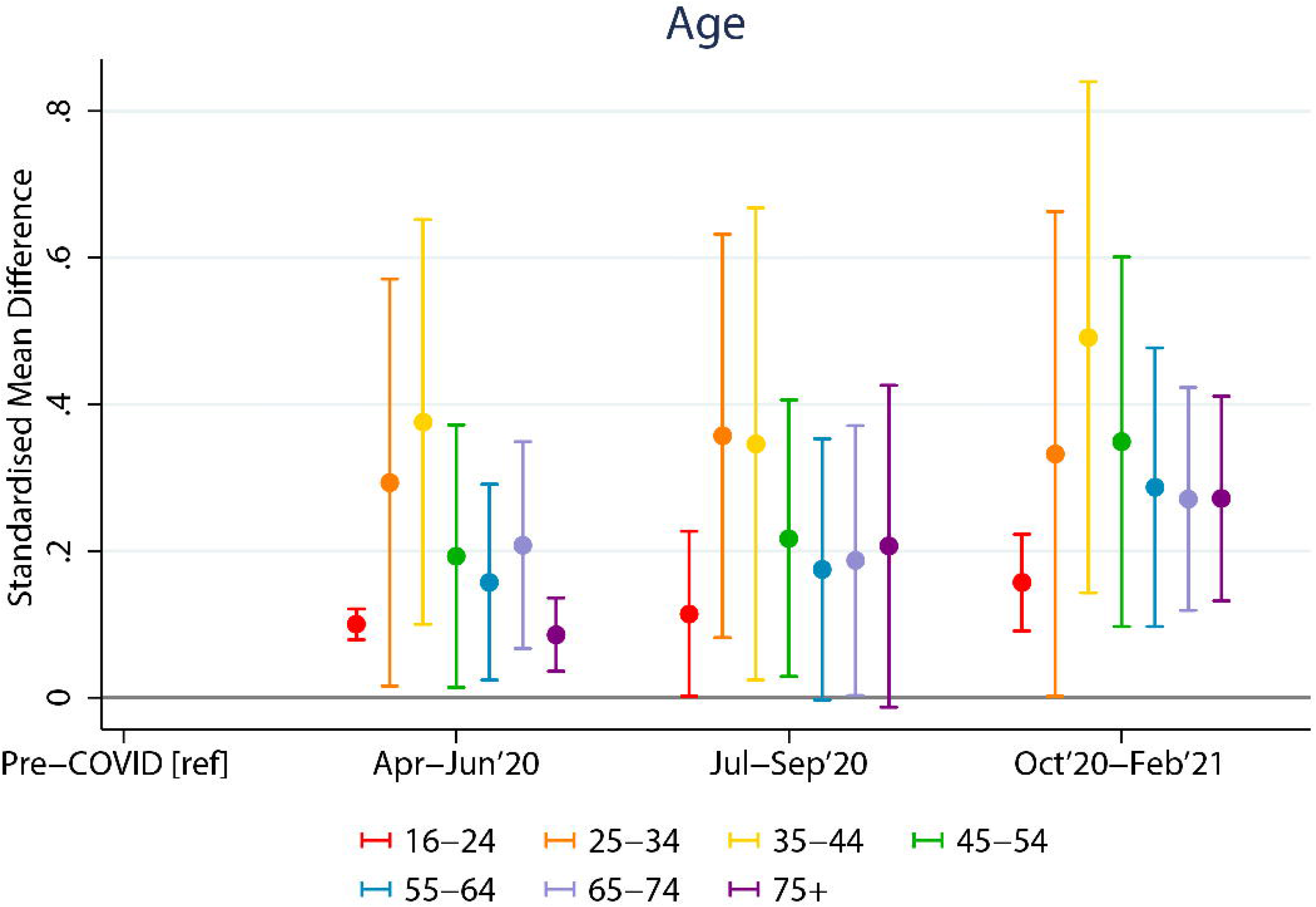
Trends in prevalence of psychological distress by age group. Stratified changes across time periods (compared to pre-pandemic distress). Each light-coloured point represents estimates from a different included study (study specific estimates can be seen in Supplement 2).

### Sensitivity and meta-regression analyses

Sensitivity analyses were conducted to consider specific measures of mental health (depression or anxiety), and to limit data to participants with survey responses during all three time periods. Findings were consistent with the main analyses (Supplement 2).

Given the high levels of heterogeneity across studies, we conducted meta-regressions to examine whether time between pre- and post-pandemic measures, measurement type, and representativeness of the studies for their target population helped account for some of the observed heterogeneity (Supplement 2). Heterogeneity was largely unexplained by these factors; the largest explanatory factor was the representativeness of the studies, which explained 3.25% of the heterogeneity at TP2 and suggested the deterioration in distress was less marked in representative studies. A subsequent meta-analysis including only studies with national coverage showed a worsening of mental health over the pandemic similar to the main meta-analysis (TP1 SMD: 0.15 (95% CI: 0.02, 0.29); TP2: 0.11 (0.05, 0.17); TP3: 0.16 (0.05, 0.27)).

## Discussion

Our analyses of 11 well-established longitudinal studies provide a comprehensive picture of the evolution of mental health over the course of differing lockdown periods during the COVID-19 pandemic. Overall, our results indicate mental health has deteriorated since the onset of the pandemic and this has been sustained with no evidence of recovery, even when lockdown measures temporarily eased in the UK during the summer of 2020. Although evidence for deterioration from pre-pandemic is seen in most included studies, there was considerable heterogeneity in effect sizes estimated. Furthermore, our findings demonstrate that while aggregate population mental health deteriorated over time, not all groups were equally affected. Women, those with a degree-level education, and young adults (aged 25-34 and 35-44) were impacted most, reporting greater increases in psychological distress during the pandemic, and thereby exacerbating some pre-pandemic mental health inequalities.

Our findings suggest that initial declines in mental health were not a transient reaction to an unprecedented event, but an early indication of a sustained deterioration from pre-pandemic levels. These findings extend research conducted earlier in the pandemic,^4,31^ replicate some research suggesting sustained effects,^9,32^ and contradict findings from some convenience samples suggesting improvements in mental health when the initial lockdown was lifted.^7^ From a policy perspective, having a wealth of longitudinal data with both pre-pandemic and during COVID-19 date gives further information on how the pandemic has impacted on mental health, beyond simple convenience sampling data. While the direct mechanisms generating poorer mental health are complex, the COVID-19 pandemic resulted in considerable economic, social, and behavioural changes, and an increase in physical co-morbidities and bereavement, therefore increased mental distress is perhaps unsurprising. Financial stressors, changes in social interactions, and disruptions to daily life may all help explain our findings.^33–36^ These results suggest that deteriorations in population mental health may be driven more by time-stable disruption and concern arising from the COVID-19 pandemic, rather than the consequences of time-specific mitigation measures such as lockdowns.

Our results highlight widening gender inequalities in mental health. Females had much higher distress levels, and showed greater deterioration during the pandemic. Possible reasons include increased childcare responsibilities that disproportionately fell to women, greater economic impacts, and reports of large increases in gender-based violence.^37^ We also observed that deterioration in lockdown periods was greater in those with degree-level education, albeit from a lower pre-pandemic level indicating educational inequalities narrowed.^31^ Our investigation of age differences show that all age groups have been adversely affected to some extent, but high psychological distress was greater in 25–44-year-olds. The mechanisms underpinning subpopulation differences remain unclear, but likely include disruptions to social interactions, changes in employment or education, and shifts in parental responsibilities and/or work life balance.^38^ For example, individuals between the age of 25 and 44 are more likely to have school-aged children and may therefore have faced additional challenges of working from home and caring for children. Moreover, younger adults have been at an increased risk of employment disruptions,^39^ as well as changes in healthy behaviours,^40,41^ which may have contributed to further deteriorations in their mental health. However, the well-documented midlife peak in psychological distress is noteworthy,^42^ and may partly explain some of the deterioration we found in these age groups.

The multiple longitudinal studies included in this paper highlight the wide range in the size of the estimated deterioration in distress from pre-pandemic levels across varied data sources that represent different populations. While we explored multiple factors (such as age, outcome measure, timing and representativeness), we could not explain much of this heterogeneity. Other factors not considered, such as rates of COVID-19 within the samples might also play a role.

## Strengths and Limitations

Our study has several strengths. By harnessing high quality existing longitudinal studies, we have robust pre-pandemic baseline data, and multiple waves of data collection capturing different time periods during the pandemic. We investigated the potential impact of COVID-19 policy responses, specifically the introduction and removal of lockdown measures. Our approach to data harmonisation allowed us to develop comparable exposure, outcome and covariate measures and pool estimates for similar time periods. Furthermore, we maximised the value of existing data by using multi-level models to include all available data. The baseline samples of many of these studies were representative of their target populations, and analyses were weighted to account for non-response. Lastly, this study combines 11 longitudinal data sources and heterogeneity between the study specific estimates was large, highlighting that documenting the results from multiple sources is more reliable for informing policy and health planning than relying on a single data source.

Despite these advantages, methodological limitations should be noted. We cannot definitively attribute changes in population mental health to the COVID-19 pandemic or related policy responses as COVID-19 was a universal exposure to everyone. However, we note that we are unaware of alternate events which would have been likely to substantially confound our analyses or their interpretation. There were differences between studies in the timing of data collection (including when pre-pandemic measures were collected) and the mental health survey instruments used, though this did not account for the high levels of statistical heterogeneity observed. Similarly, although weighting was used where possible to control for non-random response, conditioning on voluntary response may induce selection bias, as it is very plausible that the mental health of the observed differ systematically from the target population. However, the broad consistency in the direction of findings across datasets provides reassurance that the key conclusions are likely to be robust to these differences, even if the magnitude of the effect size is harder to confirm.

## Conclusions

Mental health has been persistently worse than before the pandemic, particularly among women, those with higher degrees, and 25–44-year-olds. The sustained deterioration, even when lockdown measures were eased, somewhat refutes the notion that easing lockdown measures necessarily improves mental health, and implies that there are myriad pathways leading to adverse mental health impacts. This suggests that avoiding lockdown measures alone may not maintain population mental health, and other factors should be considered. For example, health services in the UK were not able to meet their population’s mental health needs pre-pandemic, with this situation made substantially worse during the pandemic.^43^ To minimise the detrimental longer-term consequences of the pandemic, mental health care needs to encompass multiple levels of support, including investment in primary care, community mental health, and public mental health. Initiatives should target groups at greater risk of experiencing mental ill-health, including ensuring rapid access to services, but also addressing the underlying drivers of poor mental health, such as mitigating risks of unemployment, sexual violence, and poverty. Our findings highlight the need for investment in mental health support to turn the tide and improve population mental health going forward.

## Supporting information

Supplementary File 1 - Study Details and Descriptive Results

Supplementary File 2 - Regression Coefficients

Supplementary File 3 - STROBE Checklist

## Data Availability

Data for MCS (SN 8682), NS (SN 5545), BCS70 (SN 8547), NCDS (SN 6137) and all four COVID-19 surveys (SN 8658) are available through the UK Data Service.
NSHD data are available on request to the NSHD Data Sharing Committee. Interested researchers can apply to access the NSHD data via a standard application procedure. Data requests should be submitted to; further details can be found at http://www.nshd.mrc.ac.uk/data.aspx. doi:10.5522/NSHD/Q101; doi:10.5522/NSHD/Q10.
ALSPAC data is available to researchers through an online proposal system. Information regarding access can be found on the ALSPAC website (http://www.bristol.ac.uk/media-library/sites/alspac/documents/researchers/data-access/ALSPAC_Access_Policy.pdf).
All USoc data are available through the UK Data Service (SN 6614 and SN 8644).
All ELSA data are available through the UK Data Service (SN 8688 and 5050).
Access to GS data is approved by the Generation Scotland Access Committee. See https://www.ed.ac.uk/generation-scotland/for-researchers/access or for further details.
The TwinsUK Resource Executive Committee (TREC) oversees management, data sharing and collaborations involving the TwinsUK registry (for further details see https://twinsuk.ac.uk/resources-for-researchers/access-our-data/).
Data from the various BiB family studies are available to researchers; see the study website for information on how to access data (https://borninbradford.nhs.uk/research/how-to-access-data/).

## Acknowledgments

The contributing studies have been made possible because of the tireless dedication, commitment and enthusiasm of the many people who have taken part. We would like to thank the participants and the numerous team members involved in the studies including interviewers, technicians, researchers, administrators, managers, health professionals and volunteers. We are additionally grateful to our funders for their financial input and support in making this research happen.

**TwinsUK:** Claire Steves, Ruth C. E. Bowyer, Deborah Hart, María Paz García, Rachel Horsfall

**ALSPAC:** Nicholas J. Timpson, Kate Northstone, Rebecca M. Pearson

**GS:** Drew Altschul, Chloe Fawns-Ritchie, Archie Campbell, Robin Flaig

**USoc:** Michaela Benzeval

**NSHD:** Andrew Wong, Maria Popham, Karen MacKinnon, Imran Shah, Philip Curran

**MCS, NS, BCS70, NCDS:** Colleagues in survey, data, and cohort maintenance teams

**BiB:** John Wright, Dan Mason and other colleagues in cohort, survey, data maintenance teams

## Funding Acknowledgements

This work was supported by the National Core Studies, an initiative funded by UKRI, NIHR and the Health and Safety Executive. The COVID-19 Longitudinal Health and Wellbeing National Core Study was funded by the Medical Research Council (MC_PC_20059). Full funding acknowledgements for each individual study can be found as part of Supplement 1.

## Author Contributions

Conceptualisation: PP, DP, SVK, ER, ASFK, GJG, MG, EJT, EM, CLN, EF, MH, MR, GBP

Methodology: PP, SVK, GBP, ER, KP, ASFK, MG, GJG, RS, CLN, JM, CH

Analysis: KP, ER, ASFK, GJG, KW, MG, GDG, EM, EJT, CH

Initial Drafting: PP, ER, SVK, MG, ASFK, KP, GJG, KW

Data Visualisation: KP, GJG

Revising: All

Funding Acquisition: PP, DP, GBP, SVK, AS

## References

1. Holmes EA, O’Connor RC, Perry VH, et al. Multidisciplinary research priorities for the COVID-19 pandemic: a call for action for mental health science. The Lancet Psychiatry. 2020;doi:10.1016/s2215-0366(20)30168-1

2. Hotopf M, Bullmore E, O’Connor RC, Holmes EA. The scope of mental health research during the COVID-19 pandemic and its aftermath. Br J Psychiatry. Oct 2020;217(4):540–542. doi:10.1192/bjp.2020.125

3. Santomauro DF, Mantilla Herrera AM, Shadid J, et al. Global prevalence and burden of depressive and anxiety disorders in 204 countries and territories in 2020 due to the COVID-19 pandemic. The Lancet. 2021;doi:10.1016/s0140-6736(21)02143-7

4. Pierce M, Hope H, Ford T, et al. Mental health before and during the COVID-19 pandemic: a longitudinal probability sample survey of the UK population. The Lancet Psychiatry. 2020;7(10):883–892. doi:10.1016/s2215-0366(20)30308-4

5. Henderson M, Fitzsimons E, Ploubidis G, Richards M, Patalay P. Mental health during lockdown: evidence from four generations - Initial findings from the COVID-19 Survey in Five National Longitudinal Studies. London: UCL Centre for Longitudinal Studies. 2020;

6. O’Connor RC, Wetherall K, Cleare S, et al. Mental health and well-being during the COVID-19 pandemic: longitudinal analyses of adults in the UK COVID-19 Mental Health & Wellbeing study. Br J Psychiatry. Oct 21 2020:1–8. doi:10.1192/bjp.2020.212

7. Fancourt D, Steptoe A, Bu F. Trajectories of anxiety and depressive symptoms during enforced isolation due to COVID-19 in England: a longitudinal observational study. The Lancet Psychiatry. 2021;8(2):141–149. doi:10.1016/s2215-0366(20)30482-x

8. Pierce M, McManus S, Hope H, et al. Mental health responses to the COVID-19 pandemic: a latent class trajectory analysis using longitudinal UK data. The Lancet Psychiatry. 2021;8(7):610–619. doi:10.1016/s2215-0366(21)00151-6

9. Kwong ASF, Pearson RM, Smith D, Northstone K, Lawlor DA, Timpson NJ. Longitudinal evidence for persistent anxiety in young adults through COVID-19 restrictions. Wellcome Open Research. 2020;5doi:10.12688/wellcomeopenres.16206.1

10. Katikireddi SV, Hainey KJ, Beale S. The impact of COVID-19 on different population subgroups: ethnic, gender and age-related disadvantage. J R Coll Physicians Edinb. 2021;51:S40–6. doi:doi: 10.4997/JRCPE.2021.240

11. Douglas M, Katikireddi SV, Taulbut M, McKee M, McCartney G. Mitigating the wider health effects of covid-19 pandemic response. BMJ. Apr 27 2020;369:m1557. doi:10.1136/bmj.m1557

12. Blundell R, Costa Dias M, Joyce R, Xu X. COVID-19 and Inequalities. Fiscal Studies. 2020;41(2):291–319.

13. University College London. COVID-19 Longitudinal Health and Wellbeing. Accessed 14th October 2021, https://www.ucl.ac.uk/covid-19-longitudinal-health-wellbeing/

14. Di Gessa G, Maddock J, Green MJ, et al. Pre-pandemic mental health and disruptions to healthcare, economic and housing outcomes during the COVID-19 pandemic: evidence from 12 UK longitudinal studies. The British Journal of Psychiatry. 2021:1–10. doi:10.1192/bjp.2021.132

15. Joshi H, Fitzsimons E. The Millennium Cohort Study: the making of a multi-purpose resource for social science and policy. Longitudinal and Life Course Studies. 2016;7(4):409–430. doi:10.14301/llcs.v7i4.416

16. Boyd A, Golding J, Macleod J, et al. Cohort Profile: the ‘children of the 90s’--the index offspring of the Avon Longitudinal Study of Parents and Children. Int J Epidemiol. Feb 2013;42(1):111–27. doi:10.1093/ije/dys064

17. Calderwood L, Sanchez C. Next Steps (formerly known as the Longitudinal Study of Young People in England). J Open Health Data. 2016;4(e2)

18. Elliott J, Shepherd P. Cohort profile: 1970 British Birth Cohort (BCS70). Int J Epidemiol. Aug 2006;35(4):836–43. doi:10.1093/ije/dyl174

19. Power C, Elliott J. Cohort profile: 1958 British birth cohort (National Child Development Study). Int J Epidemiol. Feb 2006;35(1):34–41. doi:10.1093/ije/dyi183

20. Wadsworth M, Kuh D, Richards M, Hardy R. Cohort Profile: The 1946 National Birth Cohort (MRC National Survey of Health and Development). Int J Epidemiol. Feb 2006;35(1):49–54. doi:10.1093/ije/dyi201

21. University of Essex IfSaER, NatCen Social Research, Kantar Public. Understanding Society: Waves 1-10, 2009-2017 and Harmonised BHPS: Waves 1–18, 1991–2009 (13th edn). 2020.

22. Smith BH, Campbell A, Linksted P, et al. Cohort Profile: Generation Scotland: Scottish Family Health Study (GS:SFHS). The study, its participants and their potential for genetic research on health and illness. Int J Epidemiol. Jun 2013;42(3):689–700. doi:10.1093/ije/dys084

23. Verdi S, Abbasian G, Bowyer RCE, et al. TwinsUK: The UK Adult Twin Registry Update. Twin Res Hum Genet. Dec 2019;22(6):523–529. doi:10.1017/thg.2019.65

24. Wright J, Small N, Raynor P, et al. Cohort Profile: the Born in Bradford multi-ethnic family cohort study. Int J Epidemiol. Aug 2013;42(4):978–91. doi:10.1093/ije/dys112

25. Steptoe A, Breeze E, Banks J, Nazroo J. Cohort profile: the English longitudinal study of ageing. Int J Epidemiol. Dec 2013;42(6):1640–8. doi:10.1093/ije/dys168

26. Brown M, Goodman A, Peters A, et al. COVID-19 Survey in Five National Longitudinal Studies: Waves 1, 2 and 3 User Guide (Version 3). London: UCL Centre for Longitudinal Studies and MRC Unit for Lifelong Health and Ageing 2021;

27. Addario G, Dangerfiel P, Hussey D, Pacchiotti B, Wood M. Adapting fieldwork during the COVID-19 outbreak A methodological overview of the ELSA COVID-19 Substudy (wave 1). London: NatCen Social Research. 2020;

28. Institute for Social and Economic Research. Understanding Society COVID-19 User Guide. Version 6.0. Colchester: University of Essex. 2021;

29. Zou G. A modified poisson regression approach to prospective studies with binary data. American Journal of Epidemiology. 2004;159(7):702–706. doi:doi:10.1093/aje/kwh090

30. Higgins JP, Thompson SG. Quantifying heterogeneity in a meta-analysis. Statistics in medicine. 2002;21(11):1539–58.

31. Niedzwiedz CL, Green MJ, Benzeval M, et al. Mental health and health behaviours before and during the initial phase of the COVID-19 lockdown: longitudinal analyses of the UK Household Longitudinal Study. J Epidemiol Community Health. Sep 25 2020;doi:10.1136/jech-2020-215060

32. McGinty EE, Presskreischer R, Anderson KE, Han H, Barry CL. Psychological Distress and COVID-19-Related Stressors Reported in a Longitudinal Cohort of US Adults in April and July 2020. JAMA. Dec 22 2020;324(24):2555–2557. doi:10.1001/jama.2020.21231

33. Chandola T, Kumari M, Booker CL, Benzeval M. The mental health impact of COVID-19 and lockdown-related stressors among adults in the UK. Psychol Med. Dec 7 2020:1–10. doi:10.1017/S0033291720005048

34. Iob E, Frank P, Steptoe A, Fancourt D. Levels of Severity of Depressive Symptoms Among At-Risk Groups in the UK During the COVID-19 Pandemic. JAMA Netw Open. Oct 1 2020;3(10):e2026064. doi:10.1001/jamanetworkopen.2020.26064

35. Zheng J, Morstead T, Sin N, et al. Psychological distress in North America during COVID-19: The role of pandemic-related stressors. Social Science & Medicine. 2021;270doi:10.1016/j.socscimed.2021.113687

36. Ettman CK, Abdalla SM, Cohen GH, Sampson L, Vivier PM, Galea S. Prevalence of Depression Symptoms in US Adults Before and During the COVID-19 Pandemic. JAMA Netw Open. Sep 1 2020;3(9):e2019686. doi:10.1001/jamanetworkopen.2020.19686

37. Zhou M, Hertog E, Kolpashnikova K, Kan M-Y. Gender inequalities: Changes in income, time use and well-being before and during the UK COVID-19 lockdown. SocArXiv. 2020;doi:10.31235/osf.io/u8ytc

38. Zamarro G, Prados MJ. Gender differences in couples’ division of childcare, work and mental health during COVID-19. Review of Economics of the Household. 2021;19(1):11–40.

39. Gustafsson M. Young workers in the coronavirus crisis: Findings from the Resolution Foundation’s coronavirus survey. Accessed 12th October 2021, https://www.resolutionfoundation.org/publications/young-workers-in-the-coronavirus-crisis/

40. Bann D, Villadsen A, Maddock J, et al. Changes in the behavioural determinants of health during the COVID-19 pandemic: gender, socioeconomic and ethnic inequalities in five British cohort studies. J Epidemiol Community Health. May 26 2021;doi:10.1136/jech-2020-215664

41. Wielgoszewska B, Maddock J, Green MJ, et al. The UK Coronavirus Job Retention Scheme and changes in diet, physical activity and sleep during the COVID-19 pandemic: Evidence from eight longitudinal studies. medRxiv. 2021;doi:10.1101/2021.06.08.21258531

42. Blanchflower DG. Is happiness U-shaped everywhere? Age and subjective well-being in 145 countries. J Popul Econ. Sep 9 2020:1–50. doi:10.1007/s00148-020-00797-z

43. Rose N, Manning N, Bentall R, et al. The social underpinnings of mental distress in the time of COVID-19 - time for urgent action. Wellcome Open Res. 2020;5:166. doi:10.12688/wellcomeopenres.16123.1

44. Kessler RC, Andrews G, Colpe LJ, et al. Short screening scales to monitor population prevalences and trends in non-specific psychological distress. Psychol Med. Aug 2002;32(6):959–76. doi:10.1017/s0033291702006074

45. Angold A, Costello EJ, Messer SC, Pickles A. Development of a short questionnaire for use in epidemiological studies of depression in children and adolescents. International Journal of Methods in Psychiatric Research. 1995;5(4):237–249.

46. Goldberg D. General Health Questionnaire. NFER Publishing Company; 1978.

47. Rodgers B, Pickles A, Power C, Collishaw S, Maughan B. Validity of the Malaise Inventory in general population samples. Social psychiatry and psychiatric epidemiology. 1999;34(6):333–341.

48. Radloff LS. The CES-D scale: A self-report depression scale for research in the general population. Applied Psychological Measurement 1977;1(3):385–401.

49. Kroenke K, Spitzer RL, Williams JB. The PHQ-9: Validity of a Brief Depression Severity Measure. J Gen Intern Med. 2001;16(9):606–613. doi:doi: 10.1046/j.1525-1497.2001.016009606.x

50. Kroenke K, Strine TW, Spitzer R, Williams JBW, Berry JT, Mokdad AH. The PHQ-8 as a measure of current depression in the general population. Journal of Affective Disorders. 2009;114(1):163–173.

51. Spitzer RL, Kroenke K, Williams JBW, Lowe B. A Brief Measure for Assessing Generalized Anxiety Disorder. Arch Intern Med. 2006;166:1092–1097.

52. Zigmond AS, Snaith RP. The Hospital Anxiety and Depression Scale. Acta Psychiatrica Scandinavica. 1983;67(6):361–70.

