## Supplementary File 1 - Study Details and Descriptive Results for "Psychological Distress Before and During the COVID-19 Pandemic Among Adults in the United Kingdom: Coordinated Analyses of 11 Longitudinal Studies"

### **S1.1 Ethics and Data Access Statements For Each Study**

The most recent sweeps of the Millennium Cohort Study (**MCS**), Next Steps (**NS**), the British Cohort Study 1970 (**BCS70**), the National Child Development Study (**NCDS**) and the National Survey of Health and Development (**NSHD**) and have all been granted ethical approval by the National Health Service (NHS) Research Ethics Committee and all participants have given informed consent. Data for MCS (SN 8682), NS (SN 5545), BCS70 (SN 8547), NCDS (SN 6137) and all four COVID-19 surveys (SN 8658) are available through the UK Data Service. NSHD data are available on request to the NSHD Data Sharing Committee. Interested researchers can apply to access the NSHD data via a standard application procedure. Data requests should be submitted to; further details can be found at <http://www.nshd.mrc.ac.uk/data.aspx>. doi:10.5522/NSHD/Q101; doi:10.5522/NSHD/Q10.

Ethical approval was obtained from the Avon Longitudinal Study of Parents and Children (**ALSPAC**) Ethics and Law Committee and the Local Research Ethics Committees. The study website contains details of all the data that is available through a fully searchable data dictionary and variable search tool: <http://www.bristol.ac.uk/alspac/researchers/our-data>. ALSPAC data is available to researchers through an online proposal system. Information regarding access can be found on the ALSPAC website (<http://www.bristol.ac.uk/media-library/sites/alspac/documents/researchers/data-access/ALSPAC_Access_Policy.pdf>).

The University of Essex Ethics Committee has approved all data collection for the Understanding Society (**USoc**) main study and COVID-19 waves. No additional ethical approval was necessary for this secondary data analysis. All data are available through the UK Data Service (SN 6614 and SN 8644).

Waves 1-9 of the English Longitudinal Study of Aging (**ELSA)** were approved through the National Research Ethics Service, while the COVID-19 Sub-study was approved by the UCL Research Ethics Committee. All participants provided informed consent. All data are available through the UK Data Service (SN 8688 and 5050).

Generation Scotland (**GS**) obtained ethical approval from the East of Scotland Committee on Medical Research Ethics (on behalf of the National Health Service). Reference number 20/ES/0021. Access to data is approved by the Generation Scotland Access Committee. See <https://www.ed.ac.uk/generation-scotland/for-researchers/access> or for further details.

All waves of TwinsUK (**TwinsUK**) have received ethical approval associated with TwinsUK Biobank (19/NW/0187), TwinsUK (EC04/015) or Healthy Ageing Twin Study (H.A.T.S) (07/H0802/84) studies from NHS Research Ethics Committees at the Department of Twin Research and Genetic Epidemiology, King’s College London. The TwinsUK Resource Executive Committee (TREC) oversees management, data sharing and collaborations involving the TwinsUK registry (for further details see <https://twinsuk.ac.uk/resources-for-researchers/access-our-data/>).

Born in Bradford (**BiB**) was granted ethical approval by the National Health Service Health Research Authority Yorkshire and the Humber (Bradford Leeds) Research Ethics Committee (references: 16/YH/0320, 15/YH/0455). Data from the various BiB family studies are available to researchers; see the study website for information on how to access data (<https://borninbradford.nhs.uk/research/how-to-access-data/>).

### **S1.2 Details of Mental Health Measures in Each Longitudinal Study**

The measures used by each study are summarised in S1.3 Supplementary Tables 1 and 2.

**MCS:** The K-6^1^ is a 6-item measure of psychological distress (i.e., general anxiety and depression). Responses are rated on a 5-point Likert-type scale, and capture distress over a period of four weeks prior to administration of the scale. Scores range from 0 to 24, with a conservative cut-off of 13+ applied to indicate probable psychological distress.

**ALSPAC**: Self-reported depressive symptoms were measured using the short mood and feelings questionnaire (SMFQ).^5^ The SMFQ is a 13-item questionnaire that measures the presence of depression symptoms in the previous two weeks and was administered via postal questionnaire or in research clinics. Each item is scored between 0-2, resulting in a summed score between 0-26. Depression severity can be rated in the following score bands: 0-4 none, 5-9 mild, 10-14 moderate, 15-19 moderately severe, 20-27 severe. ALSPAC also used the seven-item Generalized Anxiety Disorder Scale (GAD-7)^11^, which is a validated, self-report measure of anxiety used widely by healthcare professionals. Individuals were asked to consider how frequently they have been bothered by a number of problems in the previous two weeks and respond on a four-point Likert scale from 0 “Not at all” to 3 “Nearly every day”. The cut-off points for mild, moderate and severe anxiety, are 5, 10, and 15 respectively, and the maximum score is 21.

**NS**: The 12-item General Health Questionnaire (GHQ)^3^ was used to detect symptoms of psychological distress in NS and USoc. The GHQ is a screening instrument designed to detect symptoms of psychological distress (i.e. general anxiety and depression). Each item is scored 0-3 resulting in scores ranging from 0-36. There is an alternative scoring where each item is scored as 0-0-1-1.

**BCS70, NCDS**: The 9-item version of the Malaise Inventory^2^ was used to assess general psychological distress. Items are scored using a simple ‘Yes/No’ response, meaning continuous scores range from 0-9. Scores of four or more are indicative of probable psychiatric distress.

**NSHD:** The General Health Questionnaire (GHQ)^3^ was also used to detect symptoms of psychological distress in NSHD. For the pre-pandemic sweep, the 28-item GHQ was used, though the 12-item GHQ was used for the subsequent 3 sweeps. A common set of 6 GHQ questions were identified and used in theses analyses. For continuous analyses, each item is scored 0-3 resulting in scores ranging from 0-18. For binary analyses, each item is scored as 0-0-1-1, resulting in scores ranging from 0-6 with a threshold of 2. Sensitivity analyses were conducted to confirm the validity of the 6-item GHQ.

**USoc**: The 12-item General Health Questionnaire (GHQ)^3^ was used, as described for NS

**ELSA**: Depressive symptoms were measured using an abbreviated 8-item version of the validated Center for Epidemiologic Studies Depression Scale (CES-D).^8^ Respondents were asked whether they had experienced any depressive symptoms, such as feeling sad or having restless sleep, in the week prior to interview. For the binary classification, we considered respondents who reported four or more depressive symptoms on the CES-D scale as having elevated depressive symptoms.

| **Pre-pandemic** | | | | **Pandemic** | | | |
| --- | --- | --- | --- | --- | --- | --- | --- |
| **Measure** | **Subscale** | **Item** | **Question** | **Measure** | **Subscale** | **Item** | **Question** |
| GHQ-28 | Somatic | 3 | Been feeling run down and out of sorts | PHQ-9 | - | 4 | Feeling tired or having little energy |
| GHQ-28 | Anxiety | 2 | Had difficulty in staying asleep once you are off | PHQ-9 | - | 3 | Trouble falling or staying asleep, or sleeping too much |
| GHQ-28 | Anxiety | 4 | Been getting edgy and bad-tempered | GAD-7 | - | 6 | Becoming easily annoyed or irritable |
| GHQ-28 | Anxiety | 5 | Been getting scared or panicky for no good reason | GAD-7 | - | 1 | Feeling nervous, anxious or on edge |
| GHQ-28 | Depression | 1 | Been thinking of yourself as a worthless person | PHQ-9 | - | 6 | Feeling bad about yourself - or that you are a failure or have let yourself or your family down |
| GHQ-28 | Depression | 4 | Thought of the possibility that you might make away with yourself | PHQ-9 | - | 9 | Thoughts that you would be better off dead or of hurting yourself in some way |
| GHQ-28 | Depression | 5 | Found at times you couldn't do anything because your nerves were too bad | PHQ-9 | - | 7 | Trouble concentrating on things, such as reading the newspaper or watching television |

**GS:** Depression and anxiety were assessed during the pandemic sweeps using the GAD-7 and the Patient Health Questionnaire (PHQ-9)^4^. The PHQ-9 is a nine-item, validated tool for the assessment of depressive symptoms experienced in the previous two weeks. Participants are asked to indicate how often they have been bothered by problems such as “Little interest or pleasure in doing things?” on a four-point Likert scale from 0 “Not at all” to 3 “Nearly every day”. The score is the sum of the nine items, to a total of 27. A score of 10 or more indicates major depression. The 28-item GHQ^3^ was used to assess pre-pandemic psychological distress, and so a comparable composite measure was created from the GAD-7 and the PHQ-9 scales to enable evaluation of change over time. Correlations between each GHQ-28 item and items of the GAD-7 and PHQ-9 scales were examined, and the text of the most highly correlated (r > 0.25) questions was then reviewed and sense-checked; matched items are presented in table S1.2.T1 below. Cut-off scores were determined using ROC curves, which found a general cut-off of >4, a depression cut-off of >5 and an anxiety cut-off of >2. The latter matched the general “proportion” of PHQ-9/GAD-7 scores required for a cut-off.

**TwinsUK**: The Hospital Anxiety and Depression Scale (HADS)^9^ is a 14-item scale used to measure levels of psychiatric distress in non-psychiatric patient populations. Responses are indicated on a 4-point ordinal Likert scale.

**BiB**: Self-reported depressive symptoms were measured using the PHQ-8^10^. The PHQ-8 is an eight-item, validated tool for the assessment of depressive symptoms experienced in the previous two weeks. Participants are asked to indicate how often they have been bothered by problems such as “Little interest or pleasure in doing things?” on a four-point Likert scale from 0 “Not at all” to 3 “Nearly every day”. The score is the sum of the eight items, to a total of 24. A score of 10 or more indicates major depression. The pre-pandemic measure was administered in a clinic setting and subsequent data collection waves were completed by post. The GAD-7 anxiety measure is also used, as described for ALSPAC.

### **S1.3 Participant Flow Diagrams**

**MCS**

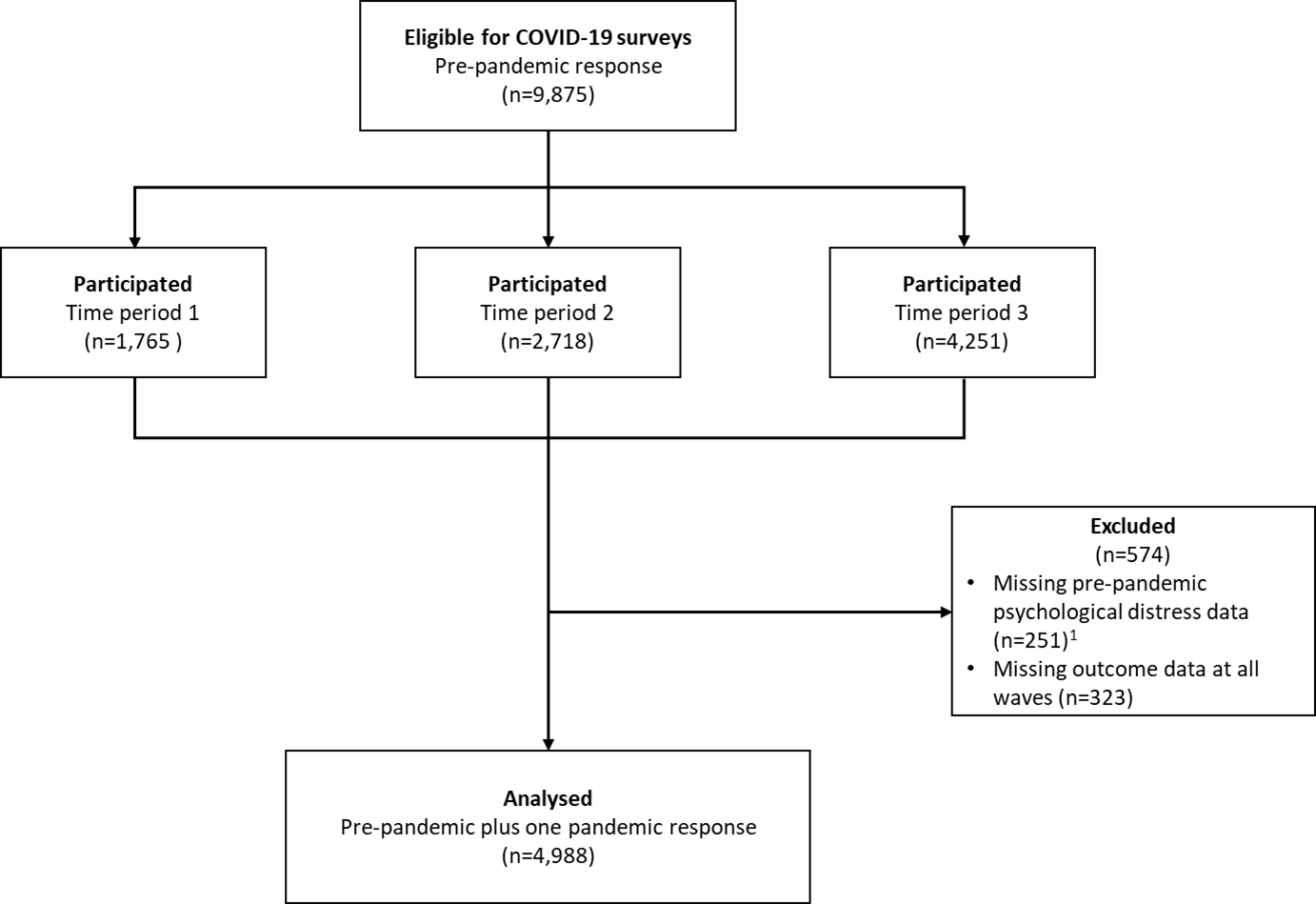

^1^ Decision to exclude from all contributing studies

**ALSPAC**

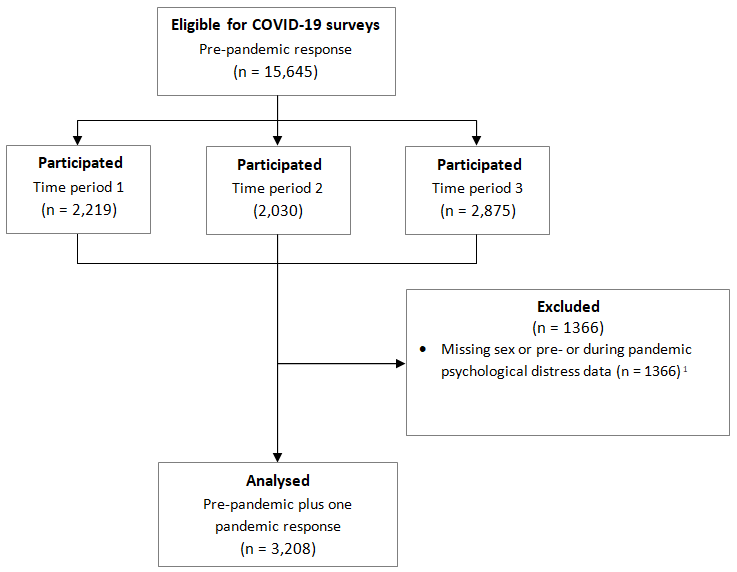

^1^ Relating to depression data only (larger sample).

**NS**

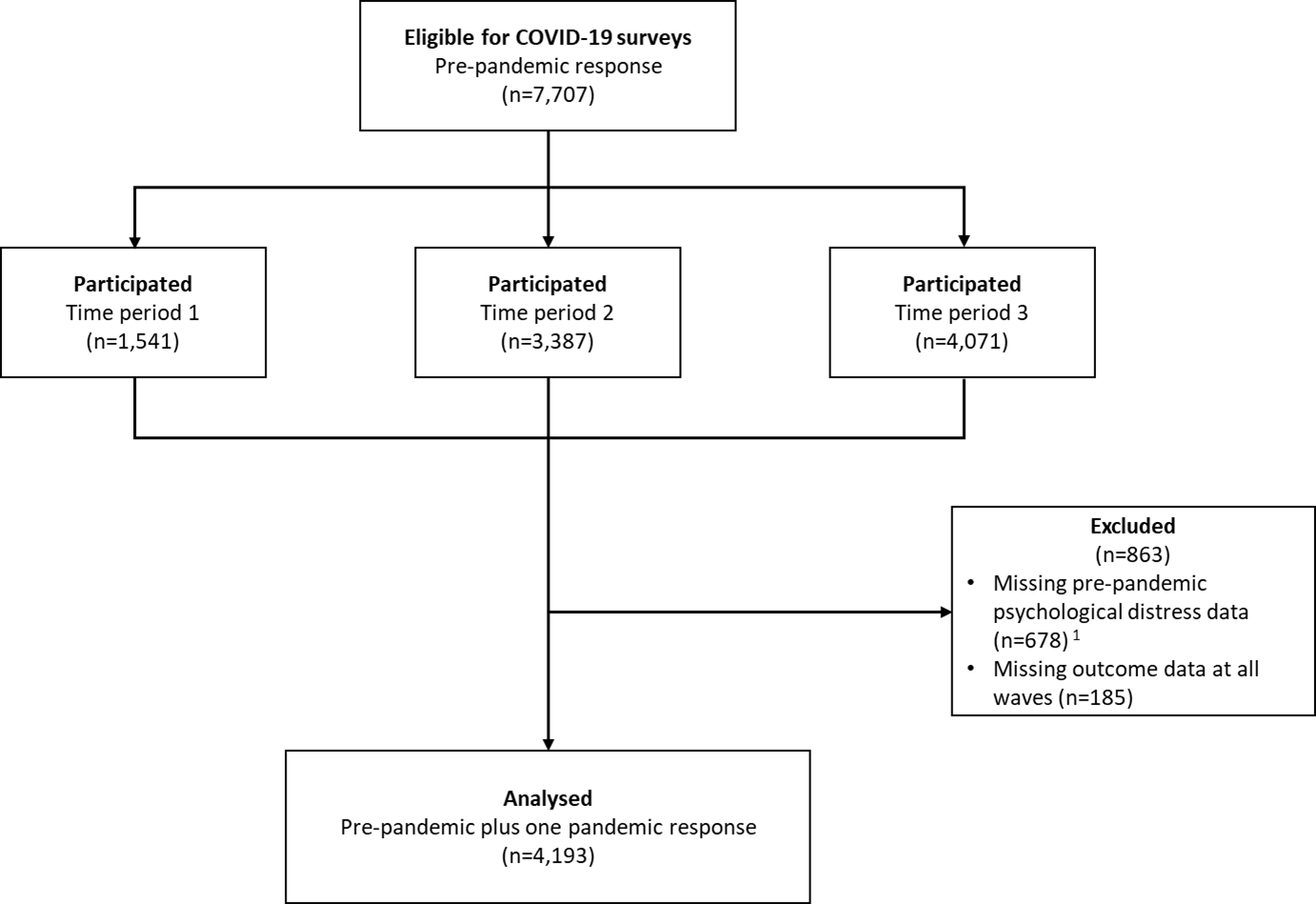

^1^ Decision to exclude from all contributing studies

**BCS70**

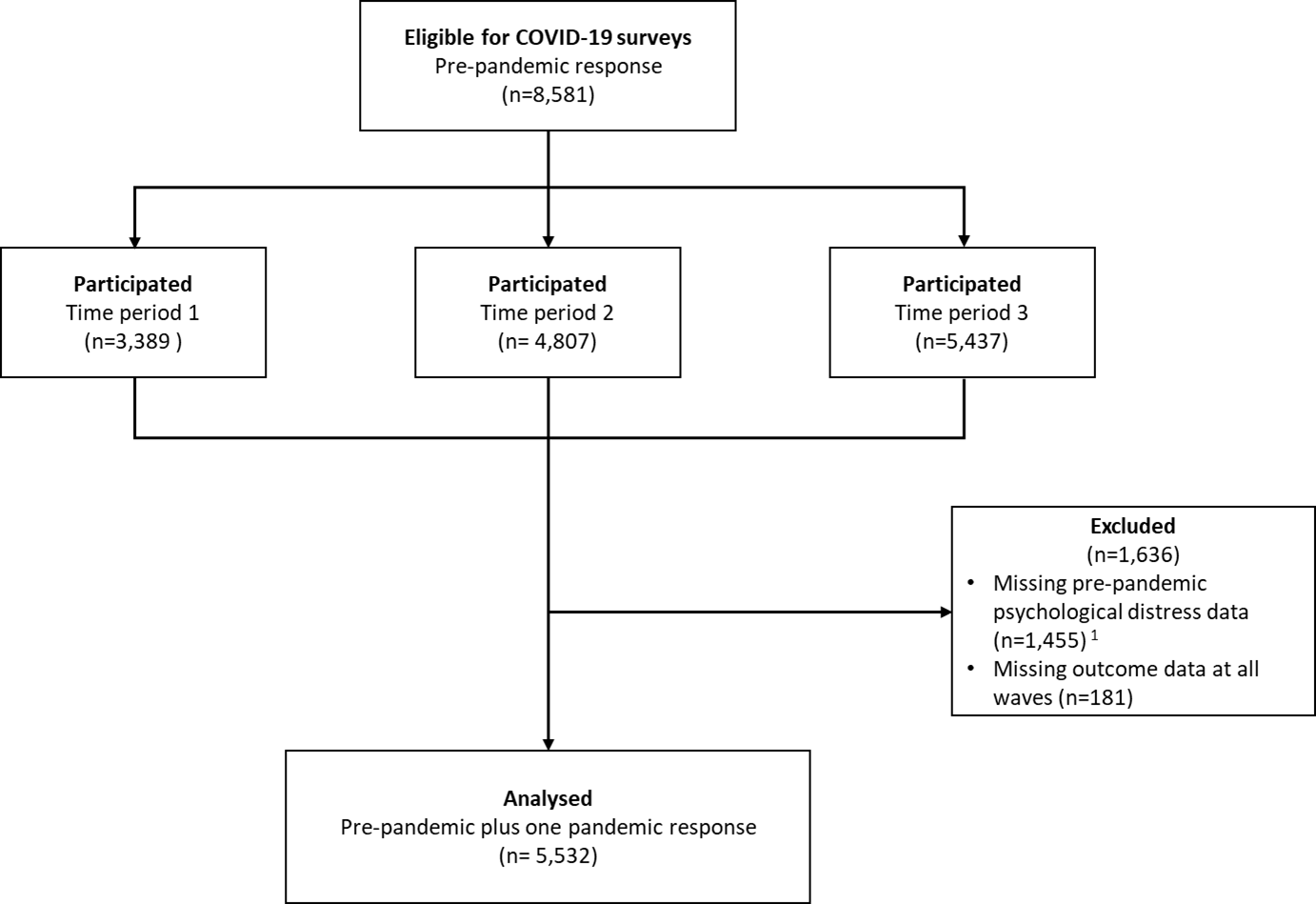

^1^ Decision to exclude from all contributing studies

**NCDS**

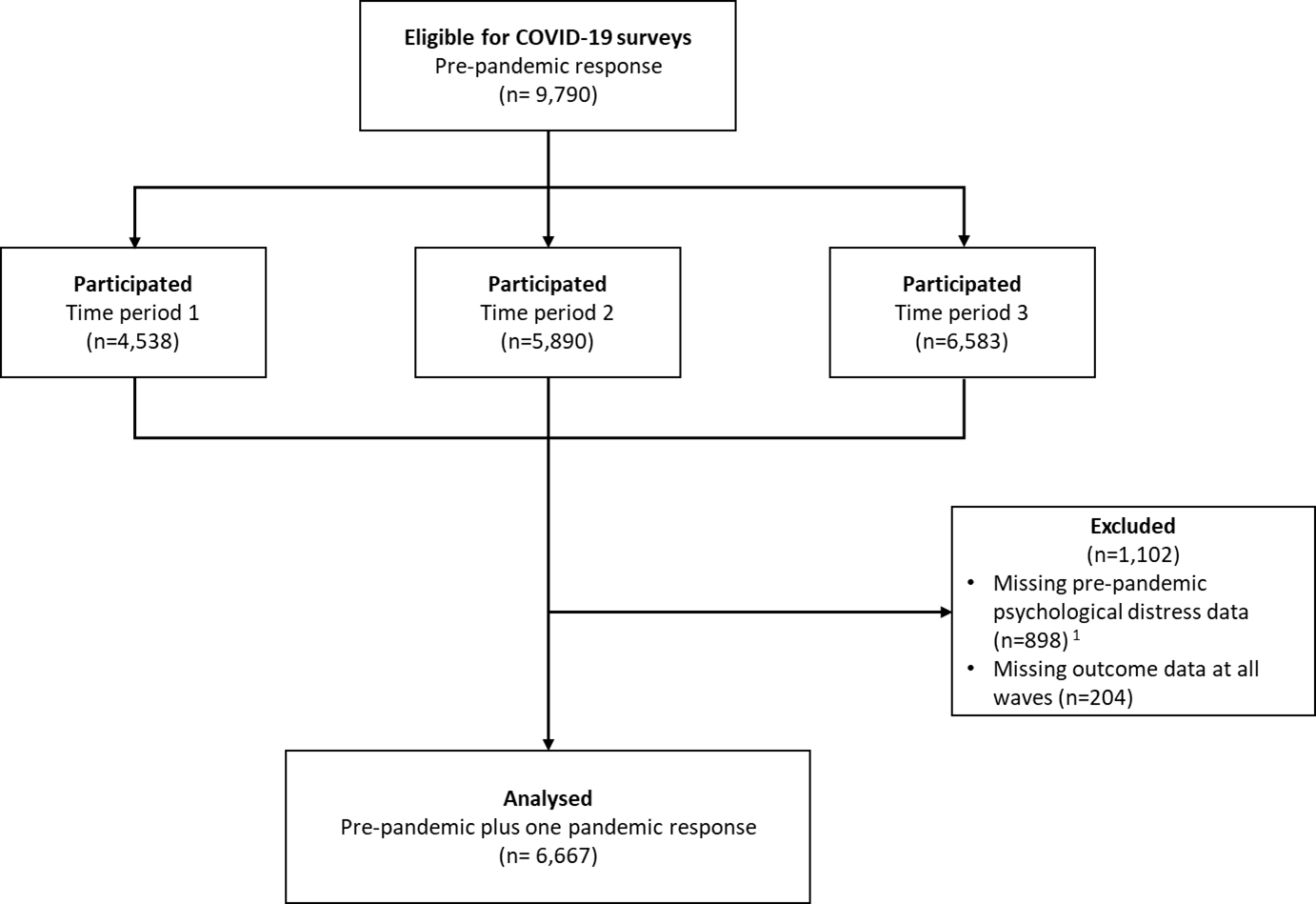

^1^ Decision to exclude from all contributing studies

**NSHD**

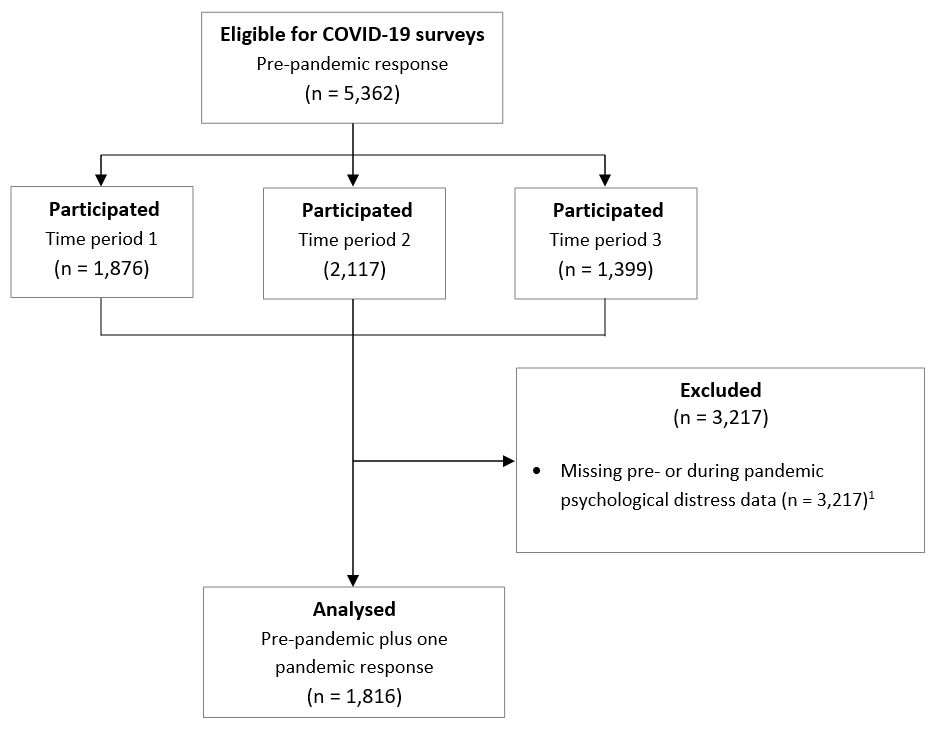

^1^ Decision to exclude from all contributing studies

**USoc**

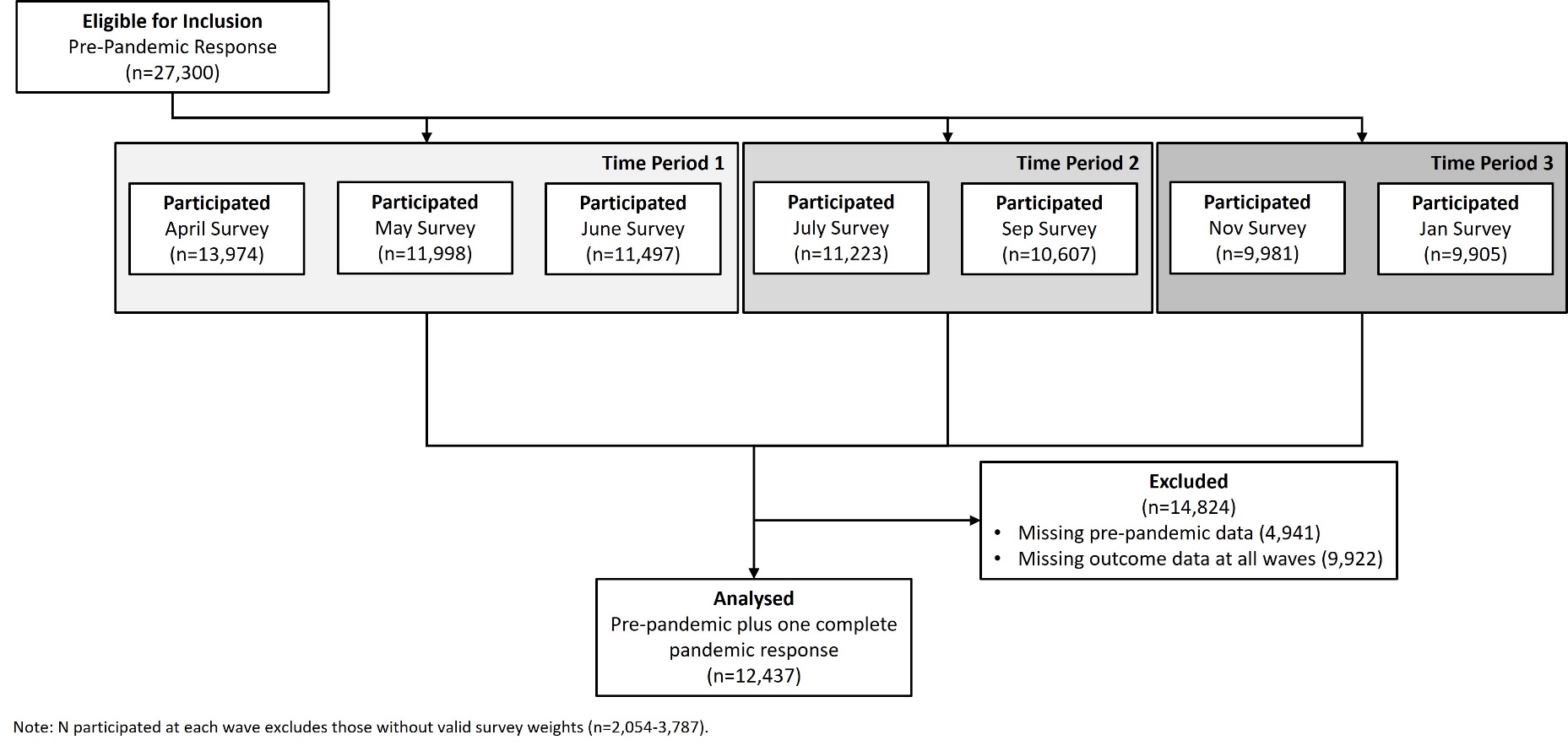

**ELSA**

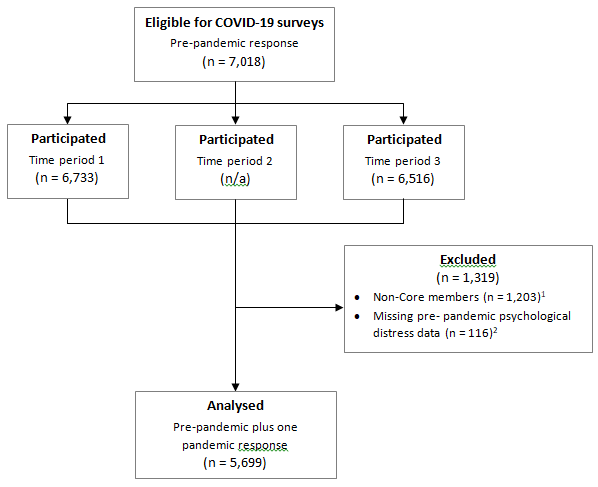

^1^ In ELSA COVID-19 Sub-Studies, longitudinal weights are produced only for ‘core’ members using ELSA wave 9 as a baseline.
 ^2^ Decision to exclude from all contributing studies

**GS**

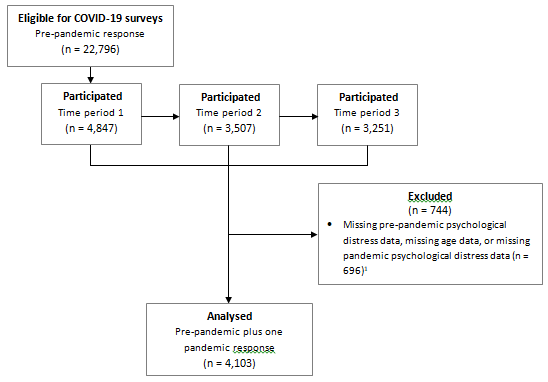

^1^ Decision to exclude from all contributing studies

**TwinsUK**

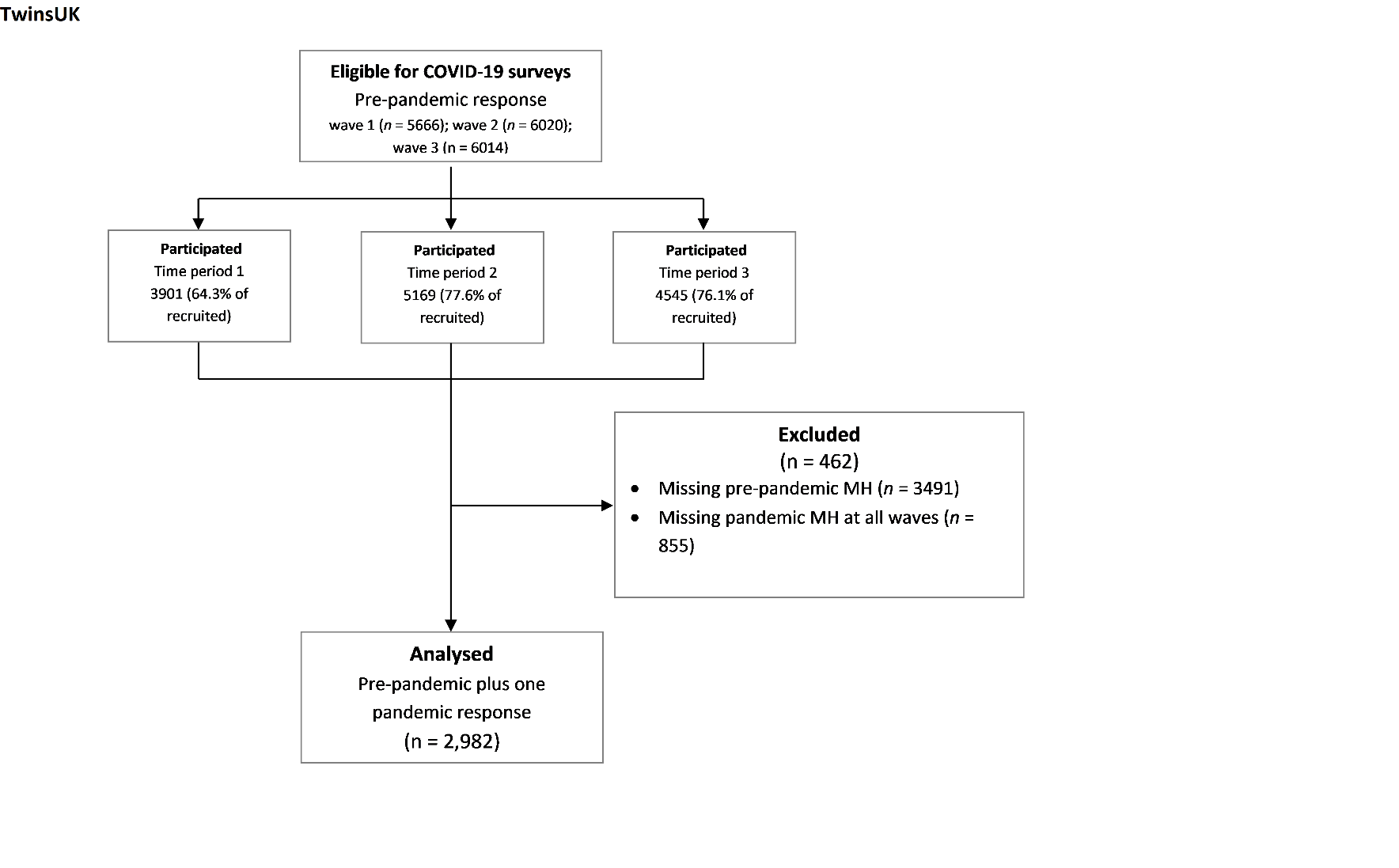

**BiB**

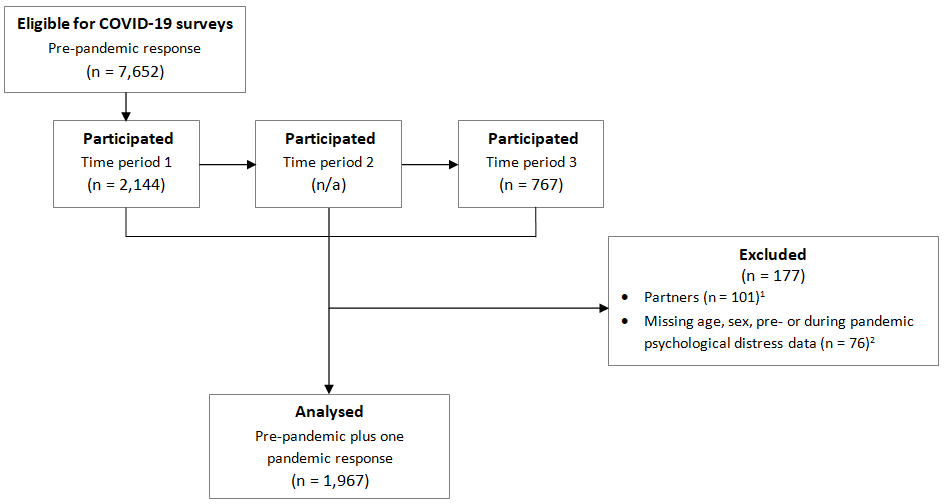

^1^ Decision to exclude made locally

^2^ Decision to exclude from all contributing studies

### **S1.3 Supplementary eTables**

**eTable 1:** Measures of general psychological distress in each study

|  | MCS | ALSPAC | NS | BCS 70 | NCDS | NSHD | USoc | ELSA | GS | TwinsUK | BiB |
| --- | --- | --- | --- | --- | --- | --- | --- | --- | --- | --- | --- |
| Measure | K-6 | SMFQ | GHQ-12 | Malaise Inventory | Malaise Inventory | GHQ-6 | GHQ-12 | CES-D-8 | Composite* | HADS | PHQ-8 |
| Number of items | 6 | 13 | 12 | 9 | 9 | 6 | 12 | 8 | 7 | 14 | 8 |
| Possible range | 0-24 | 0-26 | 0-36 | 0-9 | 0-9 | 0-18 | 0-36 | 0-70 | 0-21 | 0-36 | 0-24 |
| Threshold for high psychological distress | 13+ | 11+ | 4+ | 4+ | 4+ | 2+ | 4+ | 24+ | 5+ | 11+ | 10+ |

*see S1.2

### **eTable 2:** Additional measures of anxiety by study

|  | ALSPAC | GS | BiB |
| --- | --- | --- | --- |
| Measure | GAD-7 | Composite* | GAD-7 |
| Number of items | 7 | 2 | 7 |
| Possible range | 0-21 | 0-6 | 0-21 |
| Threshold for high psychological distress | 10+ | 3+ | 10+ |

**eTable 3:** Frequency (and %) distribution of participant characteristics by study

| **Characteristics**  (n (%)) | MCS | | ALSPAC | | NS | | BCS 70 | | NCDS | | NSHD | | USoc | | ELSA | | GS | | TwinsUK | | BiB | |
| --- | --- | --- | --- | --- | --- | --- | --- | --- | --- | --- | --- | --- | --- | --- | --- | --- | --- | --- | --- | --- | --- | --- |
| Analytic sample | 4988 | | 3208 | | 3579 | | 5532 | | 6667 | | 1816 | | 12437 | | 5699 | | 4103 | | 2981 | | 1967 | |
| **Gender** |  |  |  |  |  |  |  |  |  |  |  |  |  |  |  |  |  |  |  |  |  |  |
| Men | 1956 | (39.2) | 986 | (30.8) | 1676 | (37.4) | 2420 | (43.8) | 3145 | (47.2) | 853 | (47.0) | 5229 | (47.9) | 2440 | (47.0) | 1539 | (37.5) | 306 | (10.3) | - | - |
| Women | 3032 | (60.8) | 2219 | (69.2) | 2219 | (62.5) | 3112 | (56.3) | 3522 | (52.8) | 963 | (53.0) | 7208 | (52.1) | 3259 | (53.0) | 2565 | (62.5) | 2675 | (89.7) | 1967 | (100.0) |
| **Age group** |  |  |  |  |  |  |  |  |  |  |  |  |  |  |  |  |  |  |  |  |  |  |
| 16-24 | 4988 | (100.0) | - | - | - | - | - | - | - | - | - | - | 972 | (12.7) | - | - | - | - | 10 | (0.3) | 50 | (2.5) |
| 25-34 | - | - | 3208 | (100.0) | 4193 | (100.0) | - | - | - | - | - | - | 1278 | (12.8) | - | - | 182 | (4.4) | 89 | (3.0) | 556 | (28.2) |
| 35-44 | - | - | - | - | - | - | - | - | - | - | - | - | 1905 | (13.8) | - | - | 373 | (9.1) | 209 | (7.0) | 1006 | (51.1) |
| 45-54 | - | - | - | - | - | - | 5532 | (100.0) | - | - | - | - | 2574 | (17.2) | 642 | (18.4) | 739 | (18.0) | 323 | (10.8) | 355** | (18.0) |
| 55-64 | - | - | - | - | - | - | - | - | 6667 | (100.0) | - | - | 2641 | (17.2) | 1210 | (32.7) | 1151 | (28.1) | 654 | (21.6) | - | - |
| 65-74 | - | - | - | - | - | - | - | - | - | - | 1816 | (100.0) | 2210 | (14.7) | 2342 | (30.1) | 1393 | (33.9) | 1126 | (37.8) | - | - |
| 75+ | - | - | - | - | - | - | - | - | - | - | - | - | 857 | (11.6) | 1505 | (18.8) | 265 | (6.5) | 579 | (19.4) | - | - |
| **Education** |  |  |  |  |  |  |  |  |  |  |  |  |  |  |  |  |  |  |  |  |  |  |
| Degree | 2249 | (45.1) | 1767 | (64.7) | 2121 | (50.6) | 2983 | (53.9) | 3576 | (56.1) | 212 | (12.3) | 5917 | (37.0) | 1407 | (23.0) | 1887 | (46.0) | 1525 | (51.2) | 542 | (27.6) |
| Non-Degree | 2736 | (54.9) | 962 | (35.3) | 2072 | (49.4) | 2548 | (46.1) | 2796 | (43.9) | 1510 | (87.7) | 6520 | (63.0) | 4289 | (77.0) | 2132 | (52.0) | 1368 | (45.9) | 1013 | (51.5) |
| **Ethnicity** |  |  |  |  |  |  |  |  |  |  |  |  |  |  |  |  |  |  |  |  |  |  |
| White | 4161 | (83.8) | 2822 | (96.3) | 3073 | (74.6) | - | - | - | - | 1816 | (100.0) | 10966 | (92.5) | 5467 | (93.5) | 3994 | (96.3) | 2927 | (98.2) | 694 | (35.3) |
| Non-White | 804 | (16.2) | 108 | (3.7) | 1049 | (25.4) | - | - | - | - | - | - | 1471 | (7.5) | 232 | (6.5) | 26 | (0.6) | 47 | (1.6) | 1223 | (62.2) |
| **Country of residence** |  |  |  |  |  |  |  |  |  |  |  |  |  |  |  |  |  |  |  |  |  |  |
| England | 3165 | (82.9) | 3208 | (100.0) | 3965 | (96.8) | 4544 | (85.9) | 5453 | (84.0) | 1517 | (87.6) | 10442 | (84.8) | 5699 | (100.0) | - | - | 2733 | (91.6) | 1967 | (100.0) |
| Scotland | 578 | (9.3) | - | - | 24 | (0.6) | 426 | (8.1) | 578 | (8.9) | 138 | (8.0) | 942 | (8.0) | - | - | 4103 | (100.0) | 121 | (4.1) | - | - |
| Wales | 567 | (5.1) | - | - | 28 | (0.7) | 273 | (5.2) | 351 | (5.4) | 77 | (4.5) | 634 | (4.6) | - | - | - | - | 108 | (3.6) | - | - |
| Northern Ireland | 376 | (8.0) | - | - | 5 | (0.1) | 2 | (0.1) | 5 | (0.1) | - | - | 419 | (2.6) | - | - | - | - | 2 | (0.1) | - | - |

Notes: Samples for each study restricted to respondents with non-missing pre-pandemic psychological distress measure, with at least one response during the pandemic, and valid information on sex and age. Percentages are calculated from available data within strata. Information not available/applicable: - ; suppressed small number: *, categories collapsed e.g. Age group > 44: **

### **eTable 4:** Mean psychological distress scores and % with high psychological distress, by study and over time

|  | MCS | | | ALSPAC | | | NS | | | BCS 70 | | | NCDS | | | NSHD | | | USoc | | | ELSA | | | GS | | TwinsUK | | | BiB | | |
| --- | --- | --- | --- | --- | --- | --- | --- | --- | --- | --- | --- | --- | --- | --- | --- | --- | --- | --- | --- | --- | --- | --- | --- | --- | --- | --- | --- | --- | --- | --- | --- | --- |
| Measure | K-6 | | | SMFQ | | | GHQ-12 | | | Malaise Inventory | | | Malaise Inventory | | | GHQ-6 | | | GHQ-12 | | | CES-D-8 | | | Composite* | | HADS | | | PHQ-8 | | |
| **Mean score (SD)** |  |  | |  |  | |  |  | |  |  | |  |  | |  |  | |  |  | |  |  | |  |  |  |  | |  |  | |
| Pre-pandemic | 7.8 | | (4.9) | 6.8 | | (6.4) | 11.8 | | (6.3) | 1.9 | | (2.2) | 1.4 | | (1.9) | 4.3 | | (1.9) | 11.3 | | (5.5) | 1.3 | | (1.8) | 15.1 | (7.9) | 7.8 | | (6.2) | 4.1 | | (4.7) |
| Wave 1 | 8.0 | | (5.1) | 6.2 | | (5.6) | 13.5 | | (6.0) | 1.8 | | (2.0) | 1.4 | | (1.8) | 5.7 | | (2.8) | 12.5 | | (6.1) | 2.0 | | (2.1) | 25.4 | (4.9) | 8.6 | | (6.5) | 5.2 | | (5.4) |
| Wave 2 | 8.5 | | (5.5) | 6.5 | | (5.7) | 13.2 | | (5.7) | 2.1 | | (2.2) | 1.6 | | (2.0) | 5.4 | | (2.8) | 11.9 | | (5.7) | - | | - | 25.8 | (4.8) | 10.2 | | (5.5) | - | | - |
| Wave 3 | 9.8 | | (5.9) | 6.6 | | (6.0) | 13.4 | | (5.9) | 1.9 | | (2.1) | 1.5 | | (1.9) | 4.7 | | (1.9) | 12.9 | | (6.2) | 2.3 | | (2.4) | 24.8 | (4.7) | 10.5 | | (5.8) | 5.0 | | (5.3) |
| **High psychological distress (%)** |  |  | |  |  | |  |  | |  |  | |  |  | |  |  | |  |  | |  |  | |  |  |  |  | |  |  | |
| Pre-pandemic | 18.9 | | | 23.9 | | | 26.0 | | | 21.9 | | | 14.3 | | | 11.4 | | | 19.6 | | | 11.5 | | | 11.7 | | 6.8 | | | 11.6 | | |
| Wave 1 | 19.0 | | | 17.8 | | | 35.5 | | | 18.0 | | | 12.2 | | | 35.0 | | | 27.3 | | | 22.0 | | | 5.4 | | 7.4 | | | 18.9 | | |
| Wave 2 | 24.8 | | | 21.3 | | | 30.5 | | | 22.7 | | | 15.9 | | | 29.1 | | | 20.8 | | | - | | | 5.1 | | 6.9 | | | - | | |
| Wave 3 | 33.5 | | | 21.6 | | | 34.4 | | | 21.1 | | | 14.8 | | | 16.6 | | | 27.0 | | | 28.0 | | | 6.5 | | 7.8 | | | 16.8 | | |
| *see S1.2 | | | | | | | | | | | | | | | | | | | | | | | | | | | | | | | | |

**eTable 5:** Mean additional anxiety scores and % with psychological distress, by study and over time

*see S1.2

|  | ALSPAC | | | GS | | BiB | | |
| --- | --- | --- | --- | --- | --- | --- | --- | --- |
| Measure | GAD-7 | | | Composite* | | GAD-7 | | |
| **Mean score (SD)** |  |  | |  |  |  |  | |
| Pre-pandemic | 6.8 | | (6.4) | 0.9 | (1.0) | 3.2 | | (4.4) |
| Wave 1 | 6.2 | | (5.6) | 3.5 | (4.3) | 4.6 | | (5.2) |
| Wave 2 | 6.5 | | (5.7) | 2.8 | (4.0) | - | | - |
| Wave 3 | 6.6 | | (6.0) | 3.8 | (4.5) | 4.3 | | (5.0) |
| **High psychological distress (%)** |  |  | |  |  |  |  | |
| Pre-pandemic | 23.9 | | | 6.6 | | 10.1 | | |
| Wave 1 | 17.8 | | | 10.3 | | 15.7 | | |
| Wave 2 | 21.3 | | | 7.3 | | - | | |
| Wave 3 | 21.6 | | | 10.9 | | 13.4 | | |

**eTable 6:** Mean pre-pandemic psychological distress scores (and 95% confidence intervals) by sociodemographic characteristics and study

| **Characteristics**  (Mean (95% CI)) | MCS | ALSPAC | NS | BCS 70 | NCDS | NSHD | USoc | ELSA | GS | TwinsUK | BiB |
| --- | --- | --- | --- | --- | --- | --- | --- | --- | --- | --- | --- |
| **Overall** | 7.8 | 6.8 | 11.8 | 1.9 | 1.4 | 4.3 | 11.3 | 1.3 | 15.1 | 7.8 | 4.1 |
|  | (7.6-8.1) | (6.6-7.0) | (11.4-12.2) | (1.7-2.0) | (1.3-1.5) | (4.2-4.4) | (11.2-11.5) | (1.3-1.4) | (14.9-15.3) | (7.6-8.0) | (3.9-4.3) |
| **Gender** |  |  |  |  |  |  |  |  |  |  |  |
| Men | 6.7 | 5.2 | 11.1 | 1.5 | 1.1 | 4.0 | 10.6 | 1.0 | 14.1 | 6.8 | - |
|  | (6.3-7.1) | (4.9-5.6) | (10.5-11.8) | (1.3-1.7) | (1.0-1.2) | (3.9-4.1) | (10.4-10.8) | (1.0-1.1) | (13.7-14.5) | (6.2-7.3) |  |
| Women | 8.5 | 7.5 | 12.4 | 2.2 | 1.8 | 4.4 | 12.0 | 1.6 | 15.8 | 7.9 | 4.1 |
|  | (8.2-8.8) | (7.2-7.8) | (11.9-13.0) | (2.0-2.4) | (1.6-1.9) | (4.4-4.7) | (11.9-12.2) | (1.5-1.6) | (15.5-16.1) | (7.7-8.1) | (3.9-4.3) |
| **Age group** |  |  |  |  |  |  |  |  |  |  |  |
| 16-24 | 7.8 | - | - | - | - | - | 12.4 | - | - | 11.6 | 6.2 |
|  | (7.6-8.1) |  |  |  |  |  | (12.0-12.9) |  |  | (7.5-15.7) | (4.6-7.8) |
| 25-34 | - | 6.8 | 11.8 | - | - | - | 12.2 | - | 15.6 | 9.6 | 4.6 |
|  |  | (6.6-7.0) | (11.4-12.2) |  |  |  | (11.7-12.7) |  | (14.8-16.4) | (8.4-10.8) | (4.2-5.0) |
| 35-44 | - | - | - | - | - | - | 11.9 | - | 15.6 | 8.9 | 3.8 |
|  |  |  |  |  |  |  | (11.6-12.3) |  | (15.1-16.2) | (8.0-9.7) | (3.5-4.1) |
| 45-54 | - | - | - | 1.9 | - | - | 11.8 | 1.3 | 16.17 | 8.5 | 3.8** |
|  |  |  |  | (1.7-2.0) |  |  | (11.5-12.0) | (1.2-1.5) | (15.7-16.7) | (07.9-9.2 | (3.3-4.2) |
| 55-64 | - | - | - | - | 1.4 | - | 11.3 | 1.4 | 14.3 | 8.0 | - |
|  |  |  |  |  | (1.3-1.5) |  | (11.1-11.6) | (1.3-1.5) | (14.0-14.7) | (7.6-8.5) |  |
| 65-74 | - | - | - | - | - | 4.3  (4.2-4.4) | 9.9 | 1.2 | 12.4 | 7.1 | - |
|  |  |  |  |  |  |  | (9.7-10.1) | (1.1-1.3) | (11.6-13.2) | (6.8-7.4) |  |
| 75+ | - | - | - | - | - |  | 9.8 | 1.4 | 15.4 | 7.6 | - |
|  |  |  |  |  |  |  | (9.5-10.1) | (1.3-1.5) | (8.9-22.0) | (7.2-7.9) |  |
| **Education** |  |  |  |  |  |  |  |  |  |  |  |
| Degree | 7.7 | 6.2 | 11.5 | 1.6 | 1.2 | 4.0 | 11.2 | 1.0 | 14.9 | 7.9 | 3.5 |
|  | (7.4-8.0) | (5.9-6.5) | (11.0-12.0) | (1.5-1.7) | (1.1-1.3) | (3.8-4.2) | (11.1-11.4) | (0.9-1.1) | (14.5-15.2) | (7.6-8.2) | (3.2-3.9) |
| Non-Degree | 8.0 | 7.7 | 12.1 | 2.0 | 1.6 | 4.3 | 11.4 | 1.4 | 15.3 | 7.4 | 4.4 |
|  | (7.6-8.4) | (7.3-8.2) | (11.5-12.7) | (1.8-2.2) | (1.4-1.7) | (4.2-4.4) | (11.2-11.6) | (1.3-1.5) | (14.9-15.6) | (7.1-7.7) | (4.1-4.7) |

| **Characteristics**  (Mean (95% CI)) | MCS | ALSPAC | NS | BCS 70 | NCDS | NSHD | USoc | ELSA | GS | TwinsUK | BiB |
| --- | --- | --- | --- | --- | --- | --- | --- | --- | --- | --- | --- |
| **Ethnicity** |  |  |  |  |  |  |  |  |  |  |  |
| White | 7.9 | 6.8 | 11.7 | - | - | 4.3 | 11.3 | 1.3 | 15.1 | 9.2 | 4.3 |
|  | (7.6-8.1) | (6.6-7.0) | (11.2-12.2) |  |  | (4.2-4.4) | (11.2-11.4) | (1.2-1.3) | (14.9-15.4) | (7.4-10.9) | (3.9-4.6) |
| Non-White | 7.4 | 8.0 | 12.0 | - | - | - | 11.8 | 2.0 | 16.6 | 7.7 | 4.1 |
|  | (6.7-8.0) | (6.5-9.4) | (11.2-12.8) |  |  |  | (11.3-12.4) | (1.6-2.3) | (12.21.0) | (7.5-7.9) | (3.8-4.4) |
| **Country of residence** |  |  |  |  |  |  |  |  |  |  |  |
| England | 7.7 | 6.8 | 11.8 | 1.9 | 1.4 | 4.3 | 11.3 | 1.3 | - | 7.8 | 4.1 |
|  | (7.4-8.0) | (6.6-7.0) | (11.4-12.3) | (1.7-2.0) | (1.3-1.5) | (4.2-4.4) | (11.2-11.5) | (1.3-1.4) | - | (7.6-8.0) | (3.9-4.3) |
| Scotland | 8.2 | - | 11.3 | 2.0 | 1.3 | 4.1 | 11.6 | - | 15.1 | 7.6 | - |
|  | (7.6-8.8) |  | (7.1-15.6) | (1.6-2.4) | (0.9-1.7) | (3.9-4.4) | (11.1-12.0) |  | (14.9-15.3) | (6.5-8.7) |  |
| Wales | 8.0 | - | 9.7 | 1.7 | 1.5 | 4.2 | 11.6 | - | - | 7.7 | - |
|  | (7.4-8.6) |  | (8.0-11.4) | (1.3-2.0) | (1.1-1.9) | (3.7-4.7) | (11.1-12.2) |  |  | (6.8-8.7) |  |
| Northern Ireland | 8.2 | - | 12.5 | - | - | - | - | - | - | - | - |
|  | (7.3-9.0) |  | (-6.6-31.6) |  |  |  |  |  |  |  |  |

**Table 27:** Meta-analysed interaction coefficients

| **Modifier** | **Time Period** | **Number of Studies** | **Interaction Coefficient** | **Lower CI** | **Upper CI** | **I2** |
| --- | --- | --- | --- | --- | --- | --- |
| Sex | 1 | 10 | 0.08 | 0.03 | 0.13 | 84.93 |
| (Female) | 2 | 8 | 0.07 | 0.02 | 0.12 | 75.70 |
|  | 3 | 10 | 0.08 | 0.02 | 0.13 | 83.88 |
| Ethnicity | 1 | 7 | -0.01 | -0.13 | 0.11 | 85.80 |
| (Ethnic | 2 | 4 | 0.06 | -0.04 | 0.16 | 48.11 |
| Minority) | 3 | 7 | -0.04 | -0.16 | 0.07 | 76.78 |
| Education | 1 | 11 | -0.04 | -0.07 | -0.01 | 58.67 |
| (No Degree) | 2 | 10 | -0.01 | -0.07 | 0.06 | 88.47 |
|  | 3 | 11 | -0.05 | -0.09 | -0.01 | 67.41 |

**eTable 28:** Heterogeneity explained by measures explored by meta-regression analyses

| Measure | Timepoint | Heterogeneity (I^2^) | Heterogeneity  Explained (%) |
| --- | --- | --- | --- |
| Time between pre-pandemic and first pandemic measure | 1 | 98.22 | 1.00 |
|  | 2 | 97.78 | 0.77 |
|  | 3 | 99.17 | 1.05 |
| General population representativeness | 1 | 99.10 | 0.10 |
|  | 2 | 95.17 | 3.38 |
|  | 3 | 97.05 | 2.7 |
| Type of measure for mental health used | 1 | 98.17 | 1.05 |
|  | 2 | 95.84 | 2.19 |
|  | 3 | 98.41 | 0.83 |

**S1.4 Figures Showing Results for Binary High Psychological Distress (Caseness) Outcomes by Socio-Demographic Factor**

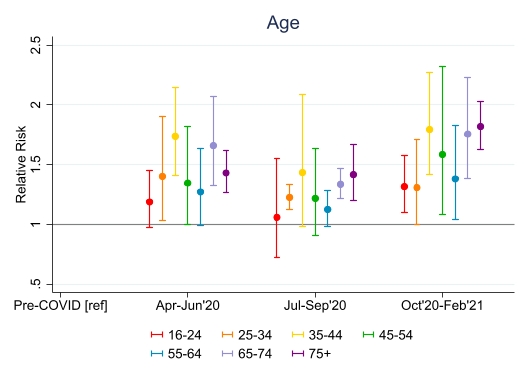

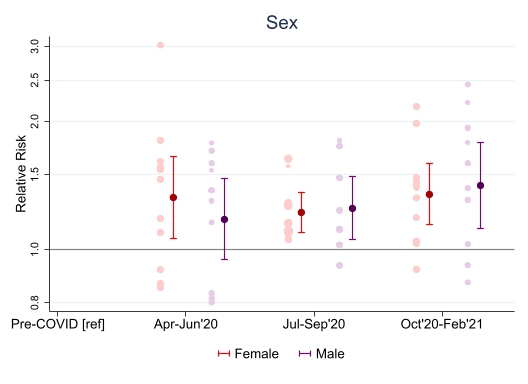

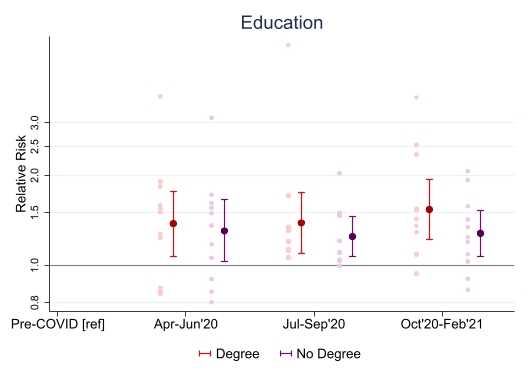

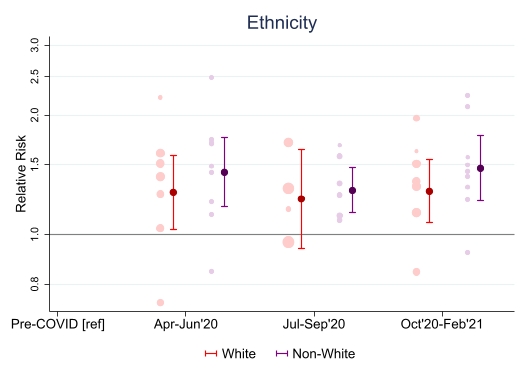

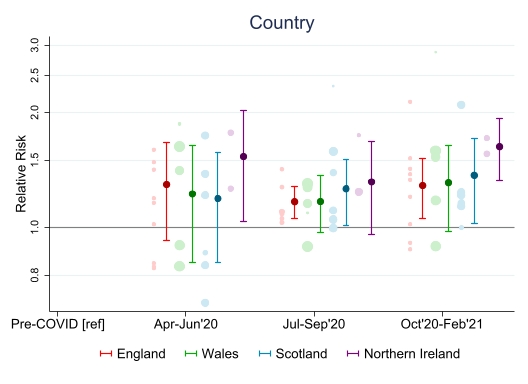

**S1.5 Meta-analysed high psychological distress when one cohort is removed**

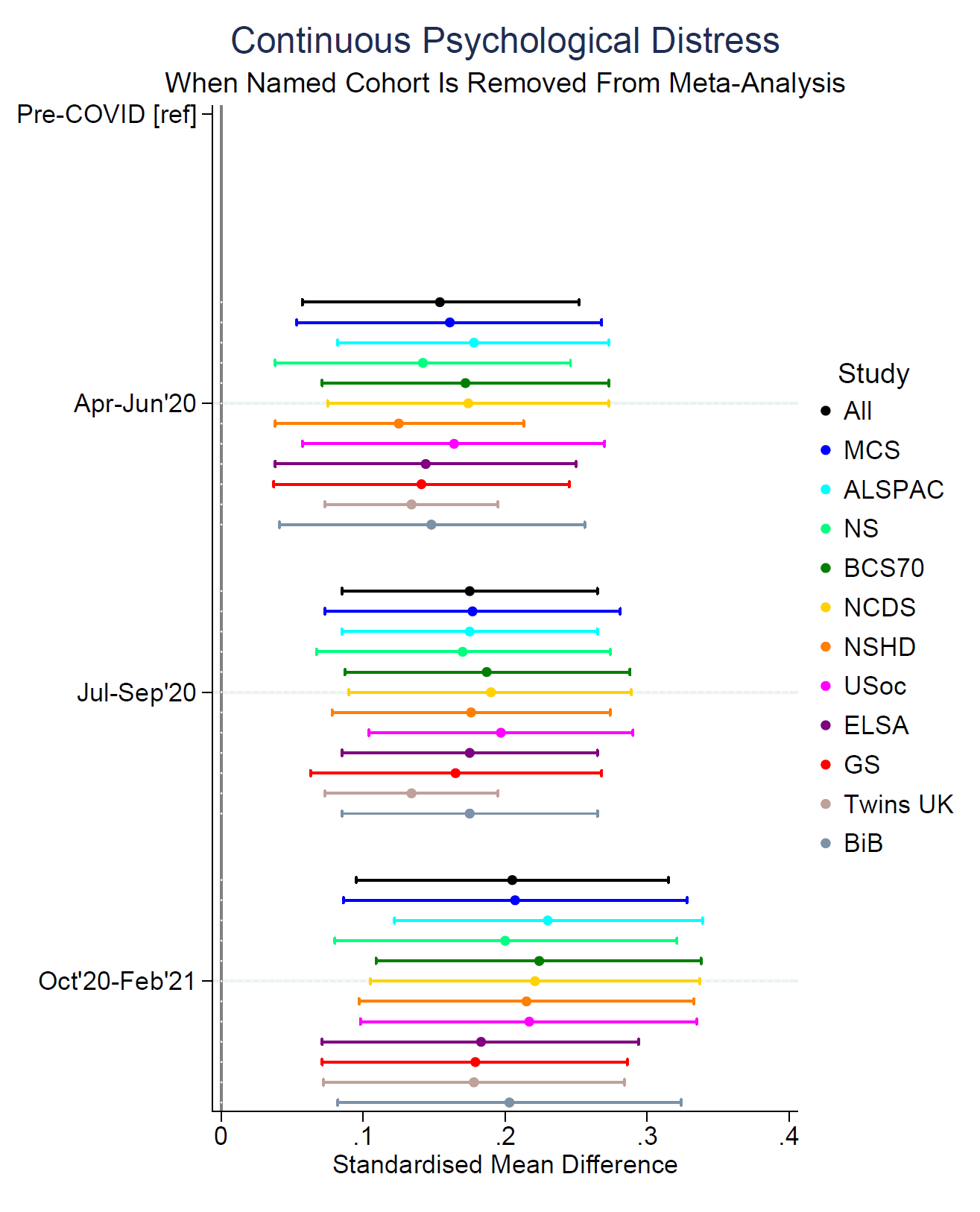

**S1.6 Funding Statements For Each Study**

**ALSPAC:** The UK Medical Research Council and Wellcome (Grant Ref: 217065/Z/19/Z) and the University of Bristol provide core support for ALSPAC. A comprehensive list of grants funding is available on the ALSPAC website (http://www.bristol.ac.uk/alspac/external/documents/grant-acknowledgements.pdf). We are extremely grateful to all the families who took part in this study, the midwives for their help in recruiting them, and the whole ALSPAC team, which includes interviewers, computer and laboratory technicians, clerical workers, research scientists, volunteers, managers, receptionists and nurses. Part of this data was collected using REDCap, see the REDCap website for details: https://projectredcap.org/resources/citations/). This work was supported by Wellcome through the Wellcome Longitudinal Population Studies COVID-19 Secretariat and Steering Group (UK LPS COVID co-ordination, Grant Ref: 221574/Z/20/Z) and supported by the Elizabeth Blackwell Institute, University of Bristol, Wellcome Trust Institutional Strategic Support Fund and Rosetrees Trust (Grant Ref: 204813/Z/16/Z; R105121). ASFK is funded by an Economics and Social Research Council (ESRC) Postdoctoral Fellowship (ES/V011650/1).

**USOC:** Understanding Society is an initiative funded by the Economic and Social Research Council and various Government Departments, with scientific leadership by the Institute for Social and Economic Research, University of Essex, and survey delivery by NatCen Social Research and Kantar Public. The Understanding Society COVID-19 study is funded by the Economic and Social Research Council (ES/K005146/1) and the Health Foundation (2076161). The research data are distributed by the UK Data Service.

**MCS, NS, BCS, NCDS, NSHD:** The Millennium Cohort Study, Next Steps, 1970 British Cohort Study and 1958 National Child Development Study are supported by the Centre for Longitudinal Studies, Resource Centre 2015-20 grant (ES/M001660/1) and a host of other co-funders. The 1946 NSHD cohort is hosted by the the MRC Unit for Lifelong Health and Ageing funded by the Medical Research Council (MC_UU_00019/1Theme 1: Cohorts and Data Collection). The COVID-19 data collections in these five cohorts were funded by the UKRI grant Understanding the economic, social and health impacts of COVID-19 using lifetime data: evidence from 5 nationally representative UK cohorts (ES/V012789/1)

**ELSA:** The English Longitudinal Study of Ageing was developed by a team of researchers based at University College London, NatCen Social Research, the Institute for Fiscal Studies, the University of Manchester and the University of East Anglia. The data were collected by NatCen Social Research. The funding is currently provided by the National Institute on Aging in the US, and a consortium of UK government departments coordinated by the National Institute for Health Research. Funding has also been received by the Economic and Social Research Council. The English Longitudinal Study of Ageing Covid-19 Substudy was supported by the UK Economic and Social Research Grant (ESRC) ES/V003941/1.

**GS:** Generation Scotland received core support from the Chief Scientist Office of the Scottish Government Health Directorates [CZD/16/6] and the Scottish Funding Council [HR03006]. Genotyping of the GS:SFHS samples was carried out by the Genetics Core   Laboratory at the Wellcome Trust Clinical Research Facility, Edinburgh, Scotland and was funded by the Medical Research Council UK and the Wellcome Trust (Wellcome Trust Strategic Award “STratifying Resilience and Depression Longitudinally” (STRADL) Reference 104036/Z/14/Z). Generation Scotland is funded by the Wellcome Trust (216767/Z/19/Z).

**TwinsUK:** TwinsUK receives funding from the Wellcome Trust (WT212904/Z/18/Z), the National Institute for Health Research (NIHR) Biomedical Research Centre based at Guy's and St Thomas' NHS Foundation Trust and King's College London. The TwinsUK COVID-19 personal experience study was funded by the King's Together Rapid COVID-19 Call award, under the projects original title ‘Keeping together through coronavirus: The physical and mental health implications of self-isolation due to the Covid-19 TwinsUK is also supported by the Chronic Disease Research Foundation and Zoe Global Ltd. The funders had no role in study design, data collection and analysis, decision to publish, or preparation of the manuscript.

**BiB:** Born in Bradford has received funding from the Wellcome Trust [101597] a joint grant from the UK Medical Research Council (MRC) and UK Economic and Social Science Research Council (ESRC) [MR/N024391/1] the National Institute for Health Research under its Applied Research Collaboration Yorkshire and Humber [NIHR200166]. This study has been funded through The Health foundation COVID-19 Award [2301201].

### **S1.7 Further Study Information**

**ALSPAC:** Pregnant women resident in Avon, UK with expected dates of delivery 1st April 1991 to 31st December 1992 were invited to take part in the study. The initial number of pregnancies enrolled is 14,541 (for these at least one questionnaire has been returned or a “Children in Focus” clinic had been attended by 19/07/99). Of these initial pregnancies, there was a total of 14,676 foetuses, resulting in 14,062 live births and 13,988 children who were alive at 1 year of age. When the oldest children were approximately 7 years of age, an attempt was made to bolster the initial sample with eligible cases who had failed to join the study originally. As a result, when considering variables collected from the age of seven onwards (and potentially abstracted from obstetric notes) there are data available for more than the 14,541 pregnancies mentioned above. The number of new pregnancies not in the initial sample (known as Phase I enrolment) that are currently represented on the built files and reflecting enrolment status at the age of 24 is 913 (456, 262 and 195 recruited during Phases II, III and IV respectively), resulting in an additional 913 children being enrolled. The phases of enrolment are described in more detail in the cohort profile paper and its update (see footnote 4 below). The total sample size for analyses using any data collected after the age of seven is therefore 15,454 pregnancies, resulting in 15,589 foetuses. Of these 14,901 were alive at 1 year of age. A 10% sample of the ALSPAC cohort, known as the Children in Focus (CiF) group, attended clinics at the University of Bristol at various time intervals between 4 to 61 months of age. The CiF group were chosen at random from the last 6 months of ALSPAC births (1432 families attended at least one clinic). Excluded were those mothers who had moved out of the area or were lost to follow-up, and those partaking in another study of infant development in Avon12.

**S1.8 References**

1. Kessler RC, Andrews G, Colpe LJ, et al. Short screening scales to monitor population prevalences and trends in non-specific psychological distress. *Psychological Medicine* 2002;32(6):959-76.

2. Rutter M, Tizard J, Whitmore K. Education, health and behaviour: Longman Publishing Group 1970.

3. Goldberg D. Manual of the general health questionnaire: Nfer Nelson 1978.

4. Kroenke K, Spitzer RL. The PHQ-9: a new depression diagnostic and severity measure. *Psychiatric Annals* 2002;32(9):509-15.

5. Angold A, Costello E, Messer S, et al. Development of a short questionnaire for use in epidemiological studies of depression in children and adolescents. *International Journal of Methods in Psychiatric Research* 1995;5:237-49.

6. Cox JL, Holden JM, Sagovsky R. Detection of postnatal depression: development of the 10-item Edinburgh Postnatal Depression Scale. *The British Journal of Psychiatry* 1987;150(6):782-86.

7. Matthey S, Barnett B, Kavanagh DJ, et al. Validation of the Edinburgh Postnatal Depression Scale for men, and comparison of item endorsement with their partners. *Journal of Affective Disorders* 2001;64(2-3):175-84.

8. Radloff LS. The CES-D scale: A self-report depression scale for research in the general population. *Applied Psychological Measurement* 1977;1(3):385-401.

9. Zigmond AS, Snaith RP. The hospital anxiety and depression scale. *Acta Psychiatrica Scandinavica* 1983;67(6):361-70.

10. Kroenke K, Strine TW, Spitzer RL, Williams JBW, Berry JT, Mokdad AH: The PHQ-8 as a measure of current depression in the general population. *Journal of Affective Disorders* 2009, 114(1):163-173.

11. Löwe B, Decker O, Müller S, Brähler E, Schellberg D, Herzog W, Herzberg PY: Validation and Standardization of the Generalized Anxiety Disorder Screener (GAD-7) in the General Population. *Medical Care* 2008, 46(3):266-274.

12. Northstone K, Lewcock M, Groom A, Boyd A, Macleod J, Timpson NJ, Wells N. The Avon Longitudinal Study of Parents and Children (ALSPAC): an updated on the enrolled sample of index children in 2019. *Wellcome Open research* 2019; 4:51.
